# Understanding COVID-19 dynamics and the effects of interventions in the Philippines: A mathematical modelling study

**DOI:** 10.1101/2021.01.14.21249848

**Authors:** Jaime M. Caldwell, Elvira de Lara-Tuprio, Timothy Robin Teng, Maria Regina Justina E. Estuar, Raymond Francis R. Sarmiento, Milinda Abayawardana, Robert Neil F. Leong, Richard T. Gray, James G. Wood, Emma S. McBryde, Romain Ragonnet, James M. Trauer

**Author notes:** corresponding author contact details: 2538 McCarthy Mall, Edmondson Hall, Honolulu, HI, 96822, USA.

## Abstract

**Objective:** COVID-19 appears to have caused less severe outbreaks in many low- and middle-income countries (LMIC) compared with high-income countries, possibly because of differing demographics, socio-economics, surveillance, and policy responses. Here, we investigate the role of multiple factors on COVID-19 dynamics in the Philippines, a LMIC that has had a relatively severe COVID-19 outbreak.

**Methods:** We applied an age-structured compartmental model that incorporated time-varying mobility, testing, and personal protective behaviors (through a “Minimum Health Standards” policy, MHS) to represent the Philippines COVID-19 epidemic nationally and for three highly affected regions (Calabarzon, Central Visayas, and the National Capital Region). We estimated effects of control measures, key epidemiological parameters, and interventions.

**Findings:** Population age structure, contact rates, mobility, testing, and MHS were sufficient to explain the Philippines epidemic based on the good fit between modelled and reported cases, hospitalisations, and deaths. Several of the fitted epidemiological parameters were consistent with those reported in high-income settings. The model indicated that MHS reduced the probability of transmission per contact by 15-26%. The February 2021 case detection rate was estimated at ∼9%, population recovered at ∼12%, and scenario projections indicated high sensitivity to MHS adherence.

**Conclusions:** COVID-19 dynamics in the Philippines are driven by age, contact structure, mobility, and MHS adherence, and the epidemic can be understood within a similar framework as for high-income settings. Continued compliance with low-cost MHS should allow the Philippines to maintain epidemic control until vaccines are widely distributed, but disease resurgence could occur due to low population immunity and detection rates.

## BACKGROUND

Coronavirus 2019 (COVID-19) epidemiology differs across settings with apparently less severe outbreaks in many low- and middle-income countries (LMIC) compared with high income countries (HIC), such that understanding why LMIC are less affected is critical to understanding global epidemiology. Multiple demographic and socio-economic factors likely drive differential COVID-19 burden across income groups. Expansive population pyramids and tropical climates may reduce COVID-19 transmission and severe disease in many LMIC (1–3), while high contact rates, high prevalence of comorbidities, high population densities, and limited healthcare capacity could increase burden (4–8). While many LMIC have had lower COVID-19 burdens than HIC, weak surveillance systems (9) could also affect epidemic estimates. Nonetheless, understanding why many LMIC have had apparently less severe outbreaks could help other countries better control their epidemics until vaccines are widely distributed.

While there are now several highly effective vaccines to prevent severe disease (10,11), and possibly infection (12,13), from SARS-CoV-2 (the disease that causes COVID-19), widespread vaccination will take months to years (14,15) and will likely need to be coupled with effective non-pharmaceutical interventions (NPIs) (16). For countries rolling out vaccines, easing NPIs too early could jeopardize success (16). However, for the majority of the world’s population, particularly those living in LMIC, vaccine access and delivery is limited (17) and widespread vaccination coverage may take two or more years (15,18). Thus, with or without vaccines, understanding the effectiveness of NPIs that can curtail COVID-19 transmission is critical. There are three broad categories of NPIs, those that aim to: (i) isolate infected individuals and their contacts; (ii) reduce contact between infected and susceptible people (henceforward “macrodistancing”); and (iii) reduce transmission given contact between infected and susceptible people (henceforward “microdistancing”). Most HIC have enforced initiatives for all three types of NPIs with a focus on long, strict macrodistancing interventions through stay-at-home orders. LMIC have also employed macrodistancing policies but often with shorter, less strict orders because these types of initiatives exacerbate poverty and have societal costs (19,20) that LMIC cannot withstand. Relaxing of short-term macrodistancing policies would be expected to lead to disease resurgence (21), which runs counter to the experience of some LMIC. A meta-analysis indicates that inexpensive microdistancing policies (e.g., wearing face masks) can be highly effective (22) and therefore could explain why some LMIC are continuing to suppress transmission after an initial period of macrodistancing.

The Philippines is one of the most severely affected countries by COVID-19 in the Western Pacific Region but has also succeeded in curtailing the epidemic while easing their most restrictive quarantine measures. Here we investigate COVID-19 epidemiology in the Philippines and the effectiveness and sensitivity to microdistancing policies. The Philippines is a LMIC that has had over 580,000 confirmed cases and more than 12,000 deaths as of 3^rd^ March 2020, with a peak in incidence in August 2020. Various NPIs have been implemented in the country, with the timing and level of restriction varying by region. The NPIs implemented include different levels of community quarantines, including school closures. Since October 2020, the Philippines has shifted from a focus on community quarantine orders towards greater reliance on Minimum Health Standards (MHS) policies, requiring the use of face coverings, physical distancing, and hand hygiene. To gain insights into the Philippines epidemic, we present a data-driven COVID-19 model, which includes age structure, heterogeneous contact patterns, time-varying testing rates, and macro- and microdistancing. We use this model to 1) estimate epidemiological parameters for COVID-19 in a LMIC for comparison with HIC; 2) consider the past effect of NPIs; and 3) create scenarios for various policy changes and estimate associated risk of disease resurgence.

## METHODS

### Model

We developed an age-structured deterministic compartmental model of SARS-CoV-2 transmission, the virus that causes COVID-19, to model disease spread at the national level, and for three highly affected regions: Calabarzon (Region IV-A), Central Visayas (Region VII), and the National Capital Region (Metro Manila). We included six sequential compartments in the model representing susceptible, non-infectious exposed, infectious exposed, early actively infectious, late actively infectious, and recovered/removed persons (Figs. 1A, S1), and stratified the infectious compartments by detection status and disease severity (Figs. 1B, S2-3; we show differential equations in supplemental material). To account for age-dependent disease processes, we stratified all model compartments by age group using 5-year age bands from birth to ≥75 years of age and allowed the proportion symptomatic, susceptibility to infection, infection fatality rate, and the probability of hospitalisation to differ by age group. We used the 2020 projected population distribution by age from the Philippines Statistics Authority for the total population sizes of each age group, which we then split among compartments (Figs. 1C, S4). To introduce heterogeneous mixing by age, we incorporated synthetic mixing matrices developed by Prem *et al*. 2017 (23) (Fig. 1D).

**Figure 1.**
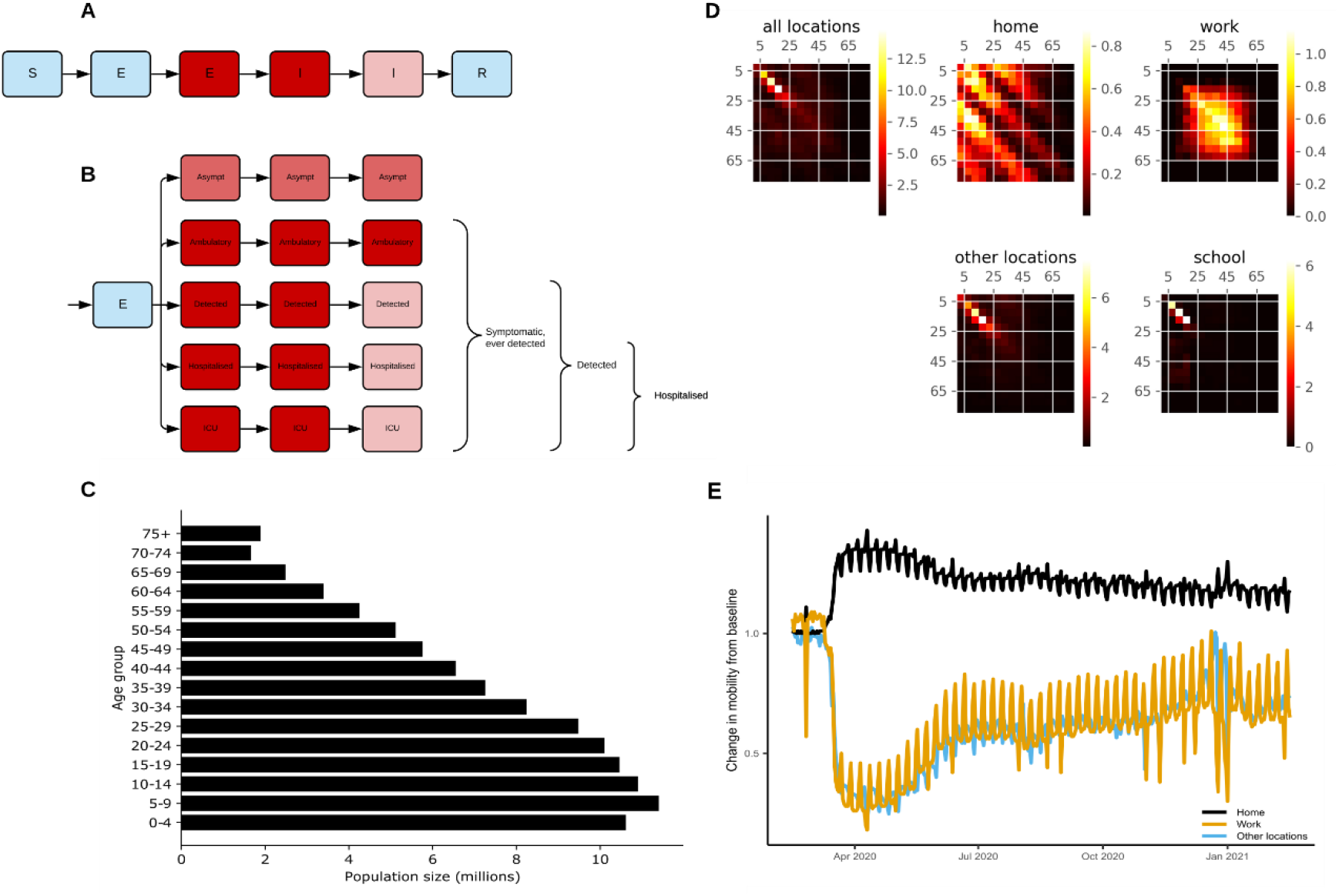
Age-structured COVID-19 model informed with population size, contact rates, and mobility from the Philippines. (A) Unstratified model structure, coloured by infectious state (blue = non-infectious; pink = moderately infectious; red = highly infectious). (B) Stratification by infection and detection status (same colour scheme as in A). (C) Starting population age distribution used in the Philippines national model. (D) Heterogeneous mixing matrices by age in the absence of NPIs (brighter colours indicate higher contact rates). (E) Community quarantine driven mobility adjustments applied to the mixing matrices (before seven-day moving average smoothing). Other locations include average from retail and recreation, supermarket and pharmacy, parks, and public transport. We provide panels C and E for the regional models in Fig. S4.

We considered several inter-compartmental transition rates to represent epidemiologically important processes. To estimate disease incidence, detected cases, hospitalisation, and ICU admission rates, we quantified transitions between different exposed and infectious compartments within the model (Fig. 1A-B, supplemental material). To calculate a modelled case detection rate, we calculated the proportion of symptomatic cases that were detected (Fig. 1B). We related the case detection rate (CDR, equation 1) to the daily number of per capita tests performed using an exponential function, under the assumption that a certain testing rate is associated with a specific case detection rate (with this parameter varied in calibration):

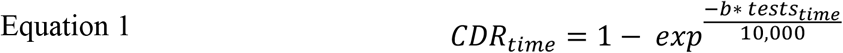

### Non-pharmaceutical interventions

We simulated community quarantines (e.g., business closures and working from home) by varying the relative contribution of four locations to an overall age-specific dynamic mixing matrix (Figs. 1E, S4). Using Google mobility data (https://www.google.com/covid19/mobility/), we scaled the household contribution to the matrix with residential mobility; the work contribution with workplace mobility; and the contribution from other locations (contacts outside of schools, homes, and work) with average mobility from all other Google mobility locations (Fig. 1E). We simulated school closures by scaling school contribution according to the proportion of children attending schools under relevant policies. In the Philippines, school closures started in mid-March 2020 in the most highly affected areas and schools remain closed throughout the country (as of 3^rd^ March 2021).

We simulated MHS as a scaled function that proportionally reduces the probability of transmission given contact (reducing the probability of an infected person passing on the infection and the probability of a contact being infected). We allowed the function to scale from zero when we assumed minimal compliance with MHS (15th August 2020) to a maximum value estimated by the calibration process (described below) when MHS was widely adopted (15th November 2020).

### Calibration

We calibrated the model to data to reproduce local COVID-19 dynamics and compared estimations of key epidemiological parameters with prior estimates from HIC (Table 1). We used three sources of local data as calibration targets: total daily confirmed cases (determined by RT-PCR) and the most recent estimate of ICU occupancy and cumulative deaths. Although we calibrated the model to these three targets simultaneously, we prioritized confirmed cases in the calibration process because these data are considered the highest quality in the Philippines for COVID-19 (based on advice from local health officials), while limiting the use of ICU and mortality data to the most recent values, as the quality of these data improved over time. To account for differences between weekday and weekend reporting, we smoothed daily confirmed cases using a seven day moving average. We simultaneously fit the model to the three calibration targets using an adaptive Metropolis algorithm (24) (Table 1; supplemental material). For the prior distributions of epidemiological calibration parameters, we used uniform priors for highly uncertain quantities and truncated normal distributions for quantities informed by epidemiological evidence (Table 1; supplemental material). To account for potential differences between HIC and LMIC, we included “adjuster” parameters to modify the proportion of symptomatic individuals, proportion of symptomatic individuals hospitalised, and the infection fatality rate as the prior distributions are based on estimates from HIC. The adjuster values are multiplicative factors applied to the odds ratio, where an adjuster value of one would indicate no adjustment is needed, a value below one would indicate the parameter is lower in the Philippines, and a value above one would indicate the parameter is higher in the Philippines. To calibrate the model, we ran seven independent chains with approximately 1000 iterations per chain and discarded the first 200 iterations as burn-in; we ensured convergence through visual inspection of trace plots. We used Latin Hypercube Sampling across the multidimensional parameter space to select initial parameter conditions. We structured the model, calibration data, likelihood function, and priors identically for the national and regional models. We report the model outputs as median values with 25-75 and 2.5-97.5 percentiles to show uncertainty.

**Table 1.**
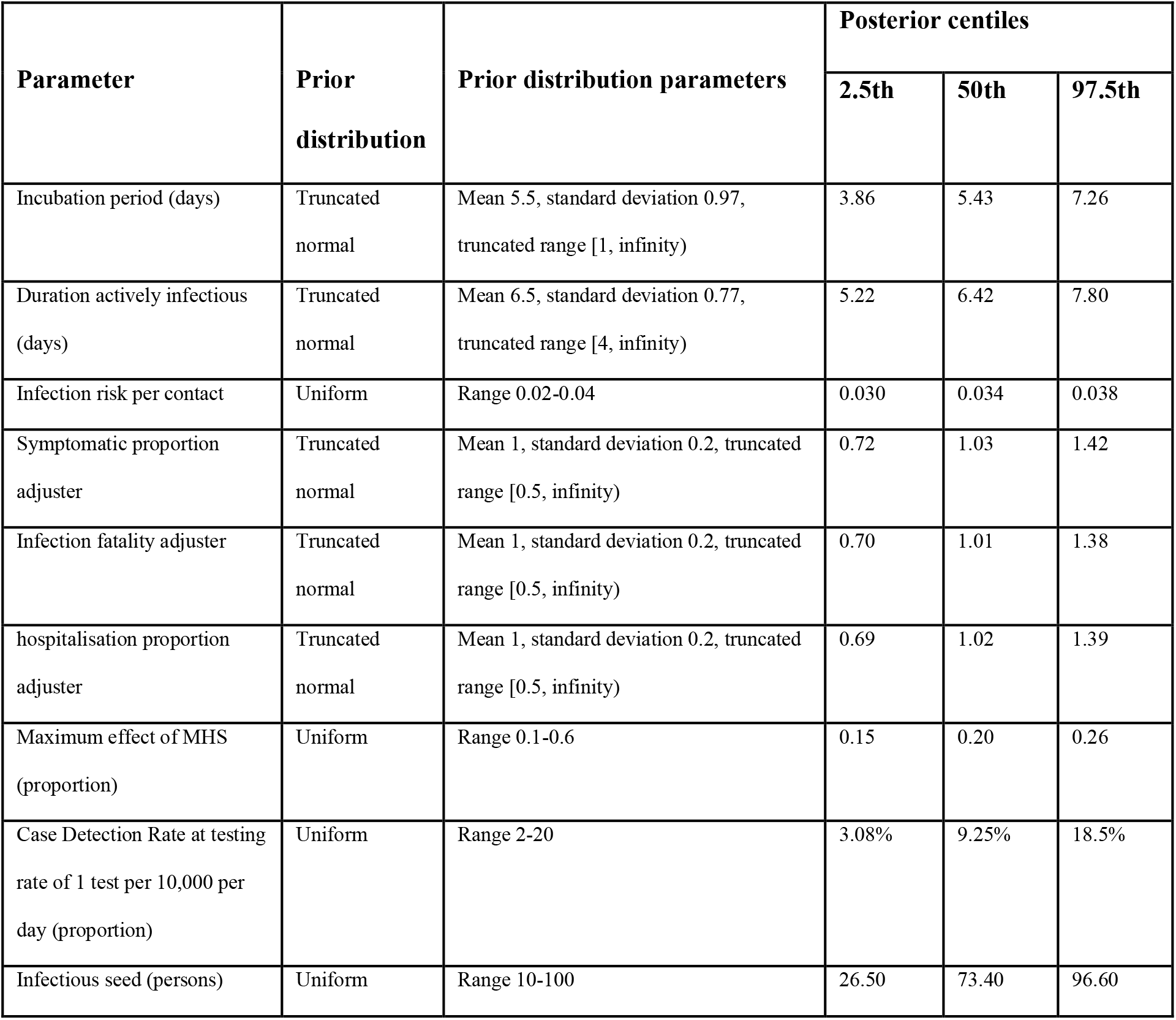
Key parameter prior and posterior distributions from the Philippines model. All parameters with the term “adjuster” allow for modification to the best estimates from the literature. Adjuster values are multiplicative factors applied to the odds ratio. An adjuster value of one indicates no adjustment is needed, a value below one indicates the parameter is lower in the Philippines, and a value above one indicates the parameter is higher in the Philippines. MHS is the Minimum Health Standards and refers to the microdistancing function that proportionally reduces the probability of transmission given contact.

| Parameter | Prior distribution | Prior distribution parameters | Posterior centiles |  |  |
| --- | --- | --- | --- | --- | --- |
|  |  |  | 2.5th | 50th | 97.5th |
| Incubation period (days) | Truncated normal | Mean 5.5, standard deviation 0.97, truncated range [1, infinity) | 3.86 | 5.43 | 7.26 |
| Duration actively infectious (days) | Truncated normal | Mean 6.5, standard deviation 0.77, truncated range [4, infinity) | 5.22 | 6.42 | 7.80 |
| Infection risk per contact | Uniform | Range 0.02-0.04 | 0.030 | 0.034 | 0.038 |
| Symptomatic proportion adjuster | Truncated normal | Mean 1, standard deviation 0.2, truncated range [0.5, infinity) | 0.72 | 1.03 | 1.42 |
| Infection fatality adjuster | Truncated normal | Mean 1, standard deviation 0.2, truncated range [0.5, infinity) | 0.70 | 1.01 | 1.38 |
| hospitalisation proportion adjuster | Truncated normal | Mean 1, standard deviation 0.2, truncated range [0.5, infinity) | 0.69 | 1.02 | 1.39 |
| Maximum effect of MHS (proportion) | Uniform | Range 0.1-0.6 | 0.15 | 0.20 | 0.26 |
| Case Detection Rate at testing rate of 1 test per 10,000 per day (proportion) | Uniform | Range 2-20 | 3.08% | 9.25% | 18.5% |
| Infectious seed (persons) | Uniform | Range 10-100 | 26.50 | 73.40 | 96.60 |

### Scenarios

We ran scenarios with different policy changes to estimate future COVID-19 transmission and potential for disease resurgence with easing different interventions. The baseline scenario held mobility and MHS constant based on the most recent date of data (26th February 2021) through the end of the scenario period (31st December 2021). All scenarios began on 27^th^ February 2021. We simulated 50% return of onsite workers by increasing the relative contribution of workplace mobility in the overall age-specific mixing matrix from current levels to current levels plus half the difference between current workplace mobility and pre- COVID workplace mobility. We simulated school reopenings by scaling school contribution in the overall age-specific mixing matrix from zero to one on the scenario start date to reflect full school attendance. To estimate the effect of easing MHS by 50, 70, and 100%, we decreased the microdistancing function, which reduces the values of all elements of the mixing matrices by a certain proportion, to 0.5, 0.3, and 0.0, respectively. We report the scenarios as median values with 25-75 and 2.5-97.5 percentiles to show uncertainty.

## RESULTS

### Epidemiological fit

The model reproduced national and regional trends in confirmed cases, hospitalisations, and deaths when incorporating changes in movement and adherence to MHS. The model fit was considerably better with the inclusion of both community quarantine-adjusted movement and adherence to MHS compared with a counterfactual scenario that only included community quarantines (Figs. 2, S5-7). These results suggest that if MHS had not been implemented, there would have likely been ten times as many cases at the outbreak peak. The four calibrated models (national and three regional models) show a reasonably good fit to total confirmed cases (Figs. 2-3). Additionally, the distributions are similar for modeled and confirmed cumulative cases by age group, with very close correspondence for those aged 40 years and above, but underpredictions for those aged 25-39 and overpredictions for ages 19 and below (Fig. S8). The models slightly overpredict reported cumulative deaths and ICU occupancy (with the exception of the National Capital Region) (Figs. 4, S9-11). We estimated an attack rate of approximately 12% (95% CI = 5-29%) as of 26th February 2021 (Fig. 4).

**Figure 2.**
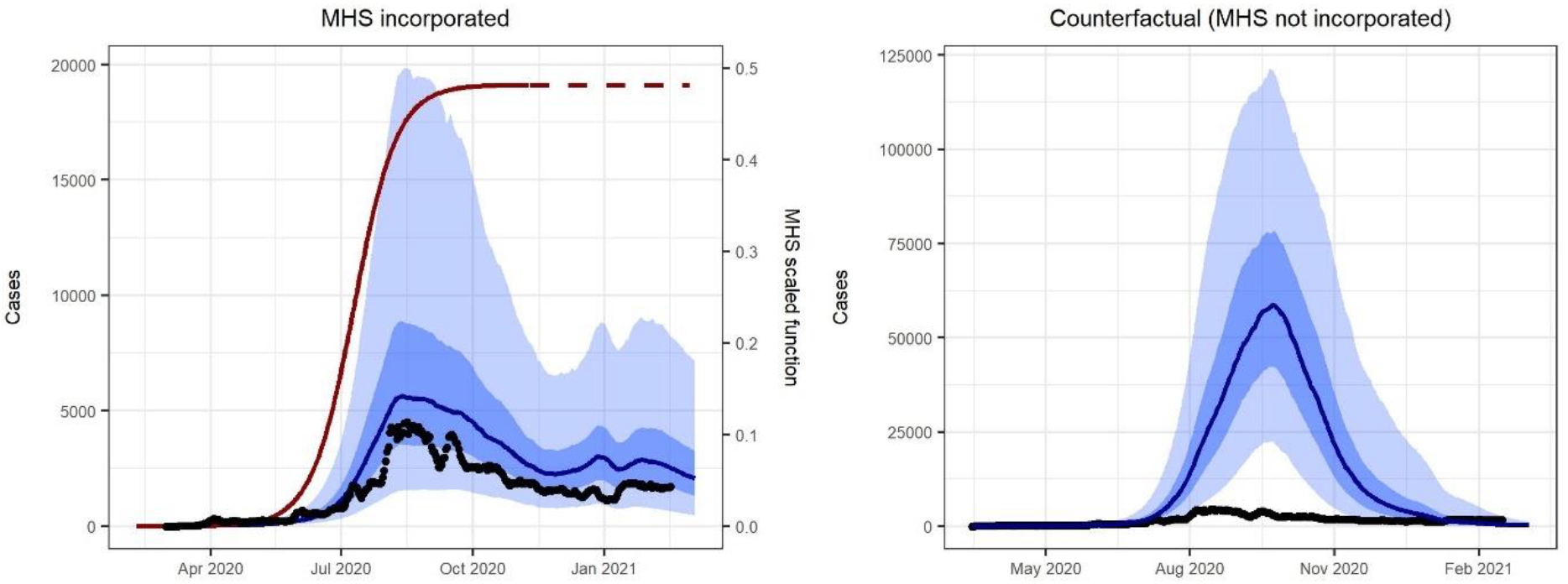
Model reproduced daily confirmed case count better with the inclusion of Minimum Health Standards (MHS). We calibrated the national model to daily confirmed cases (black dots; same in both plots with different y-axes), which included MHS (left) and ran a counterfactual scenario that did not include MHS (right). Each plot shows the median modeled detected cases (blue line) with shaded areas representing the 25th to 75th centile (dark blue) and 2.5th to 97.5th centile (light blue) of estimated detected cases. The red curve represents the effect of MHS (i.e., reduced transmission risk per contact) in the model through time. The MHS effect value is squared in the model to account for the reduction in the probability of an infected person passing on the infection and the probability of a contact being infected, prior to adjustment of each cell of the mixing matrix. We provide results for the regional models with and without MHS in Figs. S5-7.

**Figure 3:**
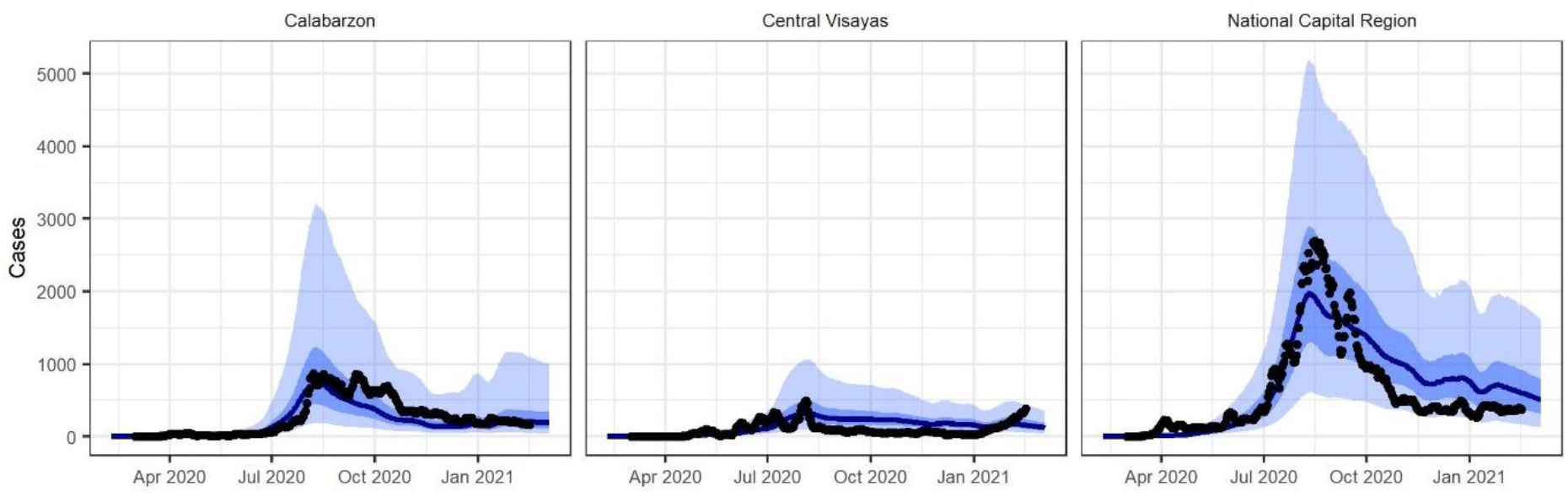
Model fit to confirmed cases in three regions of the Philippines, which varied in magnitude. Each plot shows the regional daily confirmed cases (black dots) overlaid on the median modeled detected cases (blue line), with shaded areas representing the 25th to 75th centile (dark blue) and 2.5th to 97.5th centile (light blue) of estimated detected cases.

**Figure 4.**
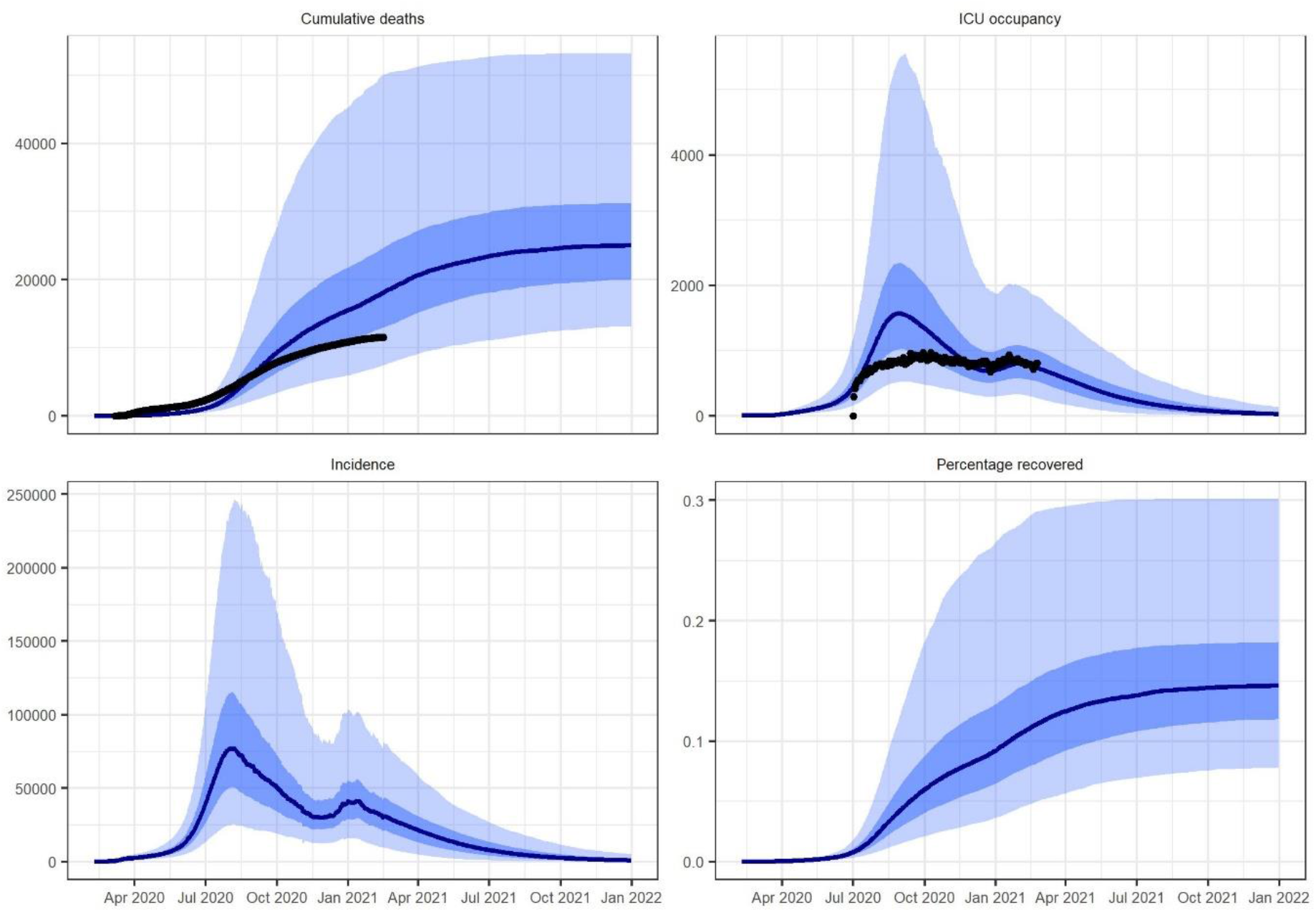
Model estimated epidemic indices from the calibrated Philippines model. Modeled median cumulative deaths, ICU occupancy, incidence, and percentage of the population recovered from COVID-19 (blue line) with shaded areas for 25th to 75th centile (dark blue) and 2.5th to 97.5th centile (light blue) and overlaid with reported cumulative deaths and ICU occupancy (black dots). Note that only the most recent estimate of cumulative deaths and ICU occupancy were included in the likelihood function, with the other time points presented as validation. ICU occupancy data was considered to have improved over the course of the epidemic. We provide equivalent regional model outputs in Figs. S9-11.

### Parameter and case detection estimation

We estimated key epidemiological parameters and transition rates (Table 1; Fig. S12). The posterior distribution of the incubation period and infectious period did not differ markedly from their prior distributions, obtained from the literature. We incorporated parameters to adjust the proportion of symptomatic individuals, the proportion of symptomatic individuals hospitalised, and the infection fatality rate from the baseline values and estimated all three adjuster parameters at approximately one, indicating no modification was needed. The maximum effect of MHS was estimated at around 0.20 (95% CI = 0.15 - 0.26) with a tight posterior distribution (Fig. S12), suggesting that MHS has reduced the risk of transmission per contact by about 20%. The model case detection was estimated at approximately 3 to 19% in the Philippines (Fig. S13), although the posterior distribution of this value was broad (Fig. S12).

### Scenario projections

We projected transmission in the near term under a range of possible scenarios and show that epidemic trajectories are highly sensitive to compliance with MHS. Moreover, allowing home workers to return onsite while maintaining MHS is estimated to have limited impact on detected cases (Figs. 5, S14-16), as well as overall incidence, hospital occupancy, and mortality. If current conditions are carried forward, transmission is projected to continue at low levels and therefore should not overwhelm hospital capacity or lead to excessive deaths. Allowing students to return to school is estimated to result in a relative high increase in cases (although this result varies markedly by region, Figs. S14-16). Reducing the effectiveness of MHS by 50% is projected to lead to a steady increase in burden, whereas reducing the effectiveness of MHS by 70% or 100% is projected to lead to a major resurgence in cases, associated hospitalisations, and deaths.

**Figure 5.**
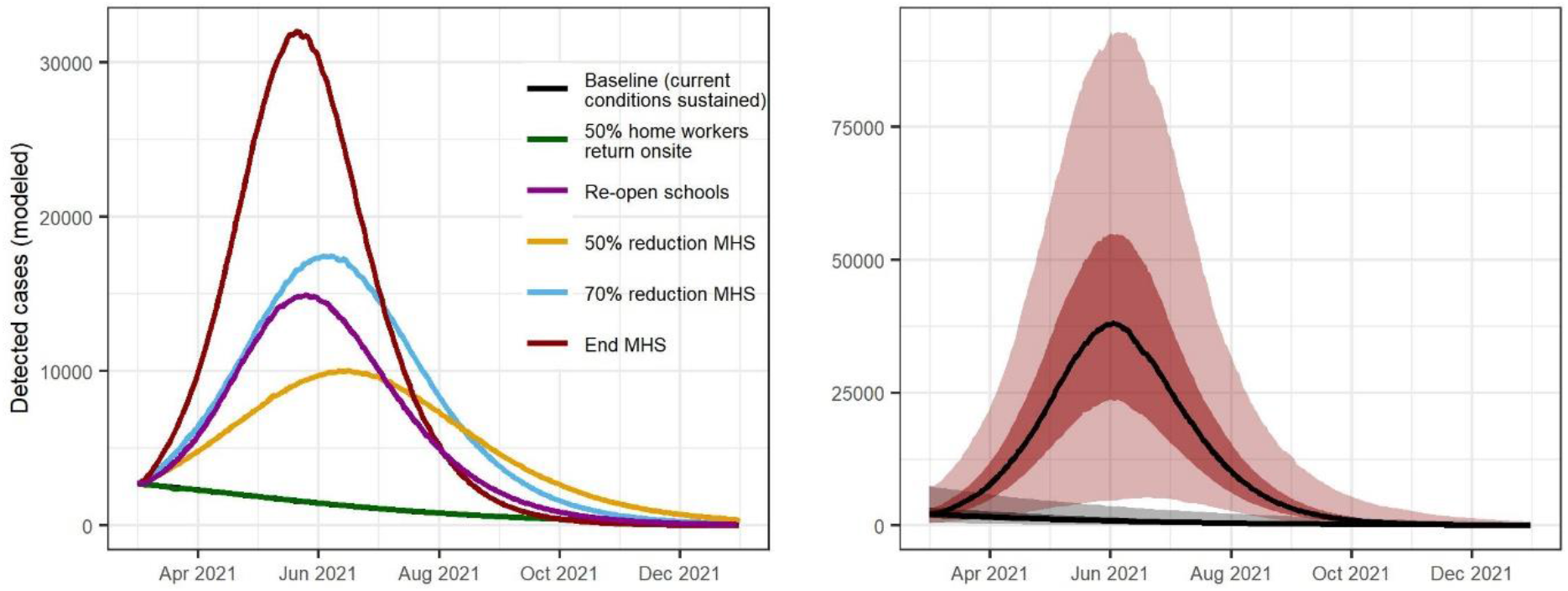
Epidemic scenario projections for detected cases from the Philippines national model, showing high sensitivity to compliance with the Minimum Health Standards (MHS). Median estimates of daily confirmed cases expected under different policy changes (left). Median estimates of daily confirmed cases (lines) with 25th to 75th centiles (dark shading) and 2.5th to 97.5th centiles (light shading) for the baseline scenario (where current conditions are carried forward) and for the scenario where MHS policy ends (right). We provide epidemic scenario projections for the regional models in Figs. S14-16.

## DISCUSSION

The combination of younger age distributions, more intergenerational contacts, changes in mobility, and changes in personal protective behaviors, appear to explain why the Philippines has sustained a relatively less severe COVID-19 outbreak than high income countries (HIC). The results from this study provide evidence that COVID-19 epidemiology in the Philippines is not inherently different from HIC. For instance, hospitalisations and infection fatality rates do not appear different from HIC, as would have been expected if healthcare capacity limitations and high prevalence of comorbidities drive disease severity and mortality rates; these findings support studies in some other LMIC (e.g., India; 18) but not others where excess deaths were considered (e.g., Brazil; 19). This highlights the importance of strong surveillance and reporting systems for understanding COVID-19 epidemiology. Younger populations typical of LMIC may reduce disease burden in two ways: 1) a proportionally smaller percent of the population is elderly (compared with HIC) and at highest risk of severe disease and mortality from COVID-19 (1,2); and 2) those who survive to old age are often of higher socioeconomic status and may have a disproportionately lower infection risk compared with the general population (27). More intergenerational contacts in LMIC may also lower disease burden because elderly people often reside with family rather than in aged-care facilities, which have driven outbreak clusters in many HIC (e.g., 21). An important finding of this study is that adherence to MHS is largely responsible for limiting transmission in the Philippines while gradually easing quarantines. These results were consistent across regions that span a wide population density gradient, suggesting that this approach could also be effective in other similar settings. These types of personal protective measures will likely continue to be needed throughout 2021 and potentially 2022, depending on the speed of vaccine distribution.

Current transmission trends in the Philippines (as of 3^rd^ March 2021) indicate that the country is maintaining low to moderate transmission, but disease resurgence remains a major threat until vaccines are widely distributed. Confirmed cases have decreased across the Philippines, plateauing at approximately 1,500-2,000 confirmed cases per day in the end of 2020 and beginning of 2021, leaving most healthcare facilities with ample ICU capacity. As people resume semi-normal activities while maintaining MHS, low levels of transmission are likely to continue, but should not overwhelm the healthcare system. The results of this study support continuing a suppression approach as a bridge to broad-scale vaccination. To support this effort, we provided real-time weekly updates of the model outputs directly to policy-makers through the Department of Health COVID-19 Philippines Local Government Units Monitoring Platform (https://fassster.ehealth.ph/covid19/). The scenarios we considered in this study indicate that relaxing societal restrictions like work-from-home orders are not predicted to cause substantial disease resurgence. Despite these findings, low case detection (11-25%) and percentage recovered (<15%) and high sensitivity to maintaining MHS could lead to disease resurgence. Limited testing is a major obstacle for the Philippines (29) and many other LMIC and can hinder our understanding of how the epidemic is progressing and associated response measures needed to prevent disease resurgence.

There are several important limitations relevant to interpreting the model results, including modelling assumptions, data limitations, and uncertainty. Most epidemiological parameters for COVID-19 used in this study were estimated from HIC (or China). While we included “adjuster” parameters to allow the symptomatic proportion, hospitalisation, and mortality rates to vary for the Philippines (similar to 25), we assumed that other parameters were similar to HIC, which may be inaccurate. However, most parameter estimates appear reasonable given previously reported COVID-19 epidemiology in the Philippines (31). When more data becomes available from the Philippines or similar settings, the current model can be updated with new parameter values. Data issues made modelling and understanding the epidemic in the Philippines particularly challenging, although data quality has progressively improved through time. For example, additional ICU facilities were created specifically for COVID-19 in the Philippines and were not required to report usage, likely affecting these estimates, especially early in the epidemic. Similarly, daily deaths, which are often considered the most reliable epidemic indicator internationally, have fluctuated markedly over the course of the epidemic, often peaking at times that appear contradictory to patterns of confirmed cases. Although reporting of daily deaths appears unreliable, cumulative deaths align reasonably well with the model projections of mortality without significant modification to the internationally estimated infection fatality rate. Finally, there is uncertainty around all model parameters, with some parameters less well constrained than others (e.g., case detection rate), which highlights the need for more epidemiological information. Future studies could build on this effort by including new, more robust data sources for model parameters and calibration targets.

## CONCLUSION

One of the most critical issues facing countries around the world is determining the best strategy for ending COVID-19 transmission through the combined use of vaccines and non-pharmaceutical interventions. More than 188 countries have been affected by COVID-19 with apparently less severe impacts in many LMIC compared with HIC. Our results suggest that COVID-19 epidemiology is similar in the Philippines, while the differences in impacts are likely driven by differences in age, contact structures, and policy, with MHS playing a substantial role in mitigating the outbreak. The Philippines has experienced one of the largest outbreaks of COVID-19 in the Western Pacific Region but also provides an approach that could help other countries relax interventions more safely, which remains an important strategy for countries with limited access to vaccines.

## Data Availability

The data analysed during the current study and model code are publicly available.

https://github.com/monash-emu/AuTuMN/tree/phl_paper_branch

## List of abbreviations

CDR: Case detection rate
COVID-19: Coronavirus disease 2019
HIC: High-income countries
ICU: Intensive care unit
LMIC: Low- and middle-income countries
MHS: Minimum Health Standards
NPI: Non-pharmaceutical intervention

## DECLARATIONS

### Ethic approval consent to participate

Not applicable.

### Consent for publication

Not applicable.

### Availability of data and materials

The data analysed during the current study and model code are publicly available at https://github.com/monash-emu/AuTuMN/tree/phl_covid_paper.

## Authors’ contributions

JMC, MA, RR, and JMT conceptualised and designed the work. JMC, EDLT, TRT, and RNFL acquired the data. JMC, EDLT, TRT, RFRS, RNFL, RTG, JGW, RR, and JMT interpreted the data. MRJEE, MA, and JMT created software used in this work. JMC drafted the manuscript and all authors substantially revised the draft. All authors approved the submitted version of the manuscript and are accountable for their own contributions to the work.

## References

1. Zhang J, Litvinova M, Liang Y, Wang Y, Wang W, Zhao S, et al. Changes in contact patterns shape the dynamics of the COVID-19 outbreak in China. Science (80-) [Internet]. 2020 Jun 26 [cited 2020 Oct 29];368(6498):1481–6. Available from: http://science.sciencemag.org/

2. O’Driscoll M, Dos Santos GR, Wang L, Cummings DAT, Azman AS, Paireau J, et al. Age-specific mortality and immunity patterns of SARS-CoV-2. Nature [Internet]. 2020 Nov 2; Available from: http://www.nature.com/articles/s41586-020-2918-0

3. Ma Y, Pei S, Shaman J, Dubrow R, Chen K. Role of air temperature and humidity in the transmission of SARS-CoV-2 in the United States 2. medRxiv [Internet]. 2020 Nov 16 [cited 2020 Nov 20];2020.11.13.20231472. Available from: https://doi.org/10.1101/2020.11.13.20231472

4. Dowd JB, Block P, Rotondi V, Mil MC. Dangerous to claim “no clear association” between intergenerational relationships and COVID-19. Proc Natl Acad Sci U S A [Internet]. 2020 Oct 20 [cited 2020 Nov 25];117(42):25975–6. Available from: www.pnas.org/cgi/doi/10.1073/pnas.2016831117

5. Brandén M, Aradhya S, Kolk M, Härkönen J, Drefahl S, Malmberg B, et al. Residential Context and COVID-19 Mortality among the Elderly in Stockholm: A population-based, observational study [Internet]. Stockholm University; 2020 [cited 2020 Nov 25]. Available from: https://cadmus.eui.eu//handle/1814/67690

6. Richardson S, Hirsch JS, Narasimhan M, Crawford JM, McGinn T, Davidson KW, et al. Presenting Characteristics, Comorbidities, and Outcomes Among 5700 Patients Hospitalized With COVID-19 in the New York City Area. JAMA [Internet]. 2020 May 26 [cited 2020 Nov 25];323(20):2052. Available from: https://jamanetwork.com/journals/jama/fullarticle/2765184

7. Rashed EA, Kodera S, Gomez-Tames J, Hirata A. Influence of Absolute Humidity, Temperature and Population Density on COVID-19 Spread and Decay Durations: Multi-Prefecture Study in Japan. Int J Environ Res Public Health [Internet]. 2020 Jul 24 [cited 2020 Nov 25];17(15):5354. Available from: https://www.mdpi.com/1660-4601/17/15/5354

8. Kadi N, Khelfaoui M. Population density, a factor in the spread of COVID-19 in Algeria: statistic study. Bull Natl Res Cent [Internet]. 2020 Dec 20 [cited 2020 Dec 2];44(1):1–7. Available from: https://link-springer-com.stanford.idm.oclc.org/articles/10.1186/s42269-020-00393-x

9. Halliday JEB, Hampson K, Hanley N, Lembo T, Sharp JP, Haydon DT, et al. Driving improvements in emerging disease surveillance through locally relevant capacity strengthening [Internet]. Vol. 357, Science. American Association for the Advancement of Science; 2017 [cited 2020 Dec 14]. p. 146–8. Available from: http://apps.who.int/iris/bitstream/

10. Oliver SE, Gargano JW, Marin M, Wallace M, Curran KG, Chamberland M, et al. The Advisory Committee on Immunization Practices’ Interim Recommendation for Use of Moderna COVID-19 Vaccine — United States, December 2020. MMWR Morb Mortal Wkly Rep [Internet]. 2021 Jan 1 [cited 2021 Mar 3];69(5152):1653–6. Available from: http://www.cdc.gov/mmwr/volumes/69/wr/mm695152e1.htm?s_cid=mm695152e1_w

11. Polack FP, Thomas SJ, Kitchin N, Absalon J, Gurtman A, Lockhart S, et al. Safety and Efficacy of the BNT162b2 mRNA Covid-19 Vaccine. N Engl J Med [Internet]. 2020 Dec 31 [cited 2021 Jan 9];383(27):2603–15. Available from: http://www.nejm.org/doi/10.1056/NEJMoa2034577

12. Dagan N, Barda N, Kepten E, Miron O, Perchik S, Katz MA, et al. BNT162b2 mRNA Covid-19 Vaccine in a Nationwide Mass Vaccination Setting. N Engl J Med [Internet]. 2021 Feb 24 [cited 2021 Mar 3];NEJMoa2101765. Available from: http://www.nejm.org/doi/10.1056/NEJMoa2101765

13. Weekes M, Jones NK, Rivett L, Workman C, Ferris M, Shaw A, et al. Single-dose BNT162b2 vaccine protects against asymptomatic SARS-CoV-2 infection. Authorea Prepr [Internet]. 2021 Feb 24 [cited 2021 Mar 3]; Available from: https://www.authorea.com/users/332778/articles/509881-single-dose-bnt162b2-vaccine-protects-against-asymptomatic-sars-cov-2-infection?access_token=-hDTQsMUXcCPSpdZV_Lmpg

14. McKee M, Rajan S. What can we learn from Israel’s rapid roll out of COVID 19 vaccination? [Internet]. Vol. 10, Israel Journal of Health Policy Research. BioMed Central Ltd; 2021 [cited 2021 Mar 3]. p. 1–4. Available from: https://link-springer-com.stanford.idm.oclc.org/articles/10.1186/s13584-021-00441-5

15. Li Bassi L. Allocating COVID-19 Vaccines Globally: An Urgent Need. JAMA Heal Forum [Internet]. 2021 Feb 8 [cited 2021 Mar 3];2(2):e210105. Available from: https://jamanetwork-com.stanford.idm.oclc.org/channels/health-forum/fullarticle/2776409

16. Gozzi NO, Bajardi P, Perra N. The importance of non-pharmaceutical interventions during the COVID-19 vaccine rollout. medRxiv [Internet]. 2021 Jan 9 [cited 2021 Mar 3];2021.01.09.21249480. Available from: https://doi.org/10.1101/2021.01.09.21249480

17. Figueroa JP, Bottazzi ME, Hotez P, Batista C, Ergonul O, Gilbert S, et al. Urgent needs of low-income and middle-income countries for COVID-19 vaccines and therapeutics. Lancet [Internet]. 2021 Feb 13 [cited 2021 Mar 3];397(10274):562–4. Available from: https://www.msf.ie/article/stigma-disrupted-care-

18. The Economist Intelligence Unit. Coronavirus vaccines: expect delays Q1 global forecast 2021. 2021.

19. Hamadani JD, Hasan MI, Baldi AJ, Hossain SJ, Shiraji S, Bhuiyan MSA, et al. Immediate impact of stay-at-home orders to control COVID-19 transmission on socioeconomic conditions, food insecurity, mental health, and intimate partner violence in Bangladeshi women and their families: an interrupted time series. Lancet Glob Heal [Internet]. 2020 Nov 1 [cited 2020 Oct 26];8(11):e1380–9. Available from: www.thelancet.com/lancetgh

20. Hogan AB, Jewell BL, Sherrard-Smith E, Vesga JF, Watson OJ, Whittaker C, et al. Potential impact of the COVID-19 pandemic on HIV, tuberculosis, and malaria in low-income and middle-income countries: a modelling study.Lancet Glob Heal [Internet]. 2020 Sep 1 [cited 2020 Oct 26];8(9):e1132–41. Available from: www.thelancet.com/lancetghVol

21. López L, Rodó X. The end of social confinement and COVID-19 re-emergence risk. Nat Hum Behav [Internet]. 2020 Jul 1 [cited 2020 Nov 25];4(7):746–55. Available from: https://doi.org/10.1038/s41562-020-0908-8

22. Haug N, Geyrhofer L, Londei A, Dervic E, Desvars-Larrive A, Loreto V, et al. Ranking the effectiveness of worldwide COVID-19 government interventions. Nat Hum Behav [Internet]. 2020 Nov 16 [cited 2020 Nov 25];1–10. Available from: https://doi.org/10.1038/s41562-020-01009-0

23. Prem K, Cook AR, Jit M. Projecting social contact matrices in 152 countries using contact surveys and demographic data. PLoS Comput Biol. 2017 Sep 1;13(9):e1005697.

24. Haario H, Saksman E, Tamminen J. An adaptive Metropolis algorithm. Bernoulli. 2001;

25. Laxminarayan R, Wahl B, Dudala SR, Gopal K, Mohan B C, Neelima S, et al. Epidemiology and transmission dynamics of COVID-19 in two Indian states. Science (80-) [Internet]. 2020 Nov 6 [cited 2020 Dec 16];370(6517):691–7. Available from: http://science.sciencemag.org/

26. Prowse TAA, Purcell T, Baía-Da-Silva DC, Sampaio V, Monteiro WM, Wood J, et al. Inferred resolution through herd immmunity of first COVID-19 wave in Manaus, Brazilian Amazon. medRxiv [Internet]. 2020 Oct 15 [cited 2020 Dec 16];2020.09.25.20201939. Available from: https://doi.org/10.1101/2020.09.25.20201939

27. Seeman TE, Crimmins E. Social environment effects on health and aging: Integrating epidemiologic and demographic approaches and perspectives. -PsycNET. Ann New York Acad Sci Popul Heal aging Strength dialogue between Epidemiol Demogr [Internet]. 2001 [cited 2020 Dec 16];954:88–117. Available from: https://psycnet-apa-org.stanford.idm.oclc.org/record/2002-00194-001

28. McMichael TM, Currie DW, Clark S, Pogosjans S, Kay M, Schwartz NG, et al. Epidemiology of Covid-19 in a Long-Term Care Facility in King County, Washington. N Engl J Med [Internet]. 2020 May 21 [cited 2020 Nov 29];382(21):2005–11. Available from: https://www-nejm-org.stanford.idm.oclc.org/doi/full/10.1056/NEJMoa2005412

29. Nuevo CE, Sigua JA, Boxshall M, Co PAW, Yap ME. Scaling Up Capacity for COVID-19 Testing in the Philippines | Coronavirus (COVID-19) Blog Posts Collection – BMJ Journals [Internet]. BMJ Journals Blog. 2020 [cited 2020 Dec 22]. Available from: https://blogs.bmj.com/covid-19/2020/06/08/scaling-up-capacity-for-covid-19-testing-in-the-philippines/

30. Pearson CAB, Kevin van Z, Jarvis CI, Davies N, Checchi F, Group CC-19 working, et al. Projections of COVID-19 epidemics in LMIC countries | CMMID Repository [Internet]. [cited 2020 May 15]. Available from: https://cmmid.github.io/topics/covid19/LMIC-projection-reports.html

31. Haw NJL, Uy J, Sy KTL, Abrigo MRM. Epidemiological profile and transmission dynamics of COVID-19 in the Philippines. Epidemiol Infect [Internet]. 2020 Sep 15 [cited 2020 Oct 29];148:e204. Available from: /core/journals/epidemiology-and-infection/article/epidemiological-profile-and-transmission-dynamics-of-covid19-in-the-philippines/FF71A51A25F004AC59885AFA4C88C48B/core-reader

