## Supplemental Materials for "Understanding COVID-19 dynamics and the effects of interventions in the Philippines: A mathematical modelling study"

Jamie M. Caldwell

Elvira de Lara-Tuprio

Timothy Robin Teng

Maria Regina Justina E. Estuar

Raymond Francis R. Sarmiento

Milinda Abayawardana

Robert Neil F. Leong

Richard T. Gray

James G. Wood

Emma S. McBryde

Romain Ragonnet

James Trauer

### 1 Base model construction

#### 1.1 Platform for infectious disease dynamics simulation

We developed a deterministic compartmental model of COVID-19 transmission using the AuTuMN platform, publicly available at https://github.com/monash-emu/AuTuMN/. Our repository allows for the rapid and robust creation and stratification of models of infectious disease epidemiology and includes pluggable modules to simulate heterogeneous population mixing, demographic processes, multiple circulating pathogen strains, repeated stratification and other modelling features relevant to infectious disease transmission. The platform was created to simulate TB dynamics, being an infectious disease whose epidemiology differs markedly by setting, such that considerable flexibility is desirable [1]. We have progressively developed the structures of our platform over recent years, and further adapted it to be sufficiently flexible to permit simulation of other infectious diseases for the purpose of this project.

#### 1.2 Base COVID-19 model

Using the base framework of an SEIR model (susceptible, exposed, infectious, removed), we split the exposed and infectious compartments into two sequential compartments each (SEEIIR). The two sequential exposed compartments represent the non-infectious and infectious phases of the incubation period, with the latter representing the "presymptomatic" phase such that infectiousness occurs during three of the six sequential phases. For this reason, "active" is a more accurate term for the two sequential "I" compartments and is preferred henceforward. The two infectious compartments represent early and late phases of active disease, during which symptoms occur if the disease episode is symptomatic, and allow explicit representation of notification, case isolation, hospitalisation and admission to ICU. The "active" compartment also includes some persons who remain asymptomatic throughout their disease episode, such that these compartments does not map directly to either persons who are infectious or those who are symptomatic (Figure 1).

The latently infected and infectious presymptomatic periods together comprise the incubation period, with the incubation period and the proportion of this period for which patients are infectious defined by input parameters described below. In general, two sequential compartments can be used to form a gamma-distributed profile of transition to infectiousness following exposure if the progression rates for these two compartments are equal, although in implementing this model the relative sojourn times in the two sequential compartments usually differed. Nevertheless, the profiles implemented are broadly consistent with the empirically observed log-normal distribution of individual incubation periods [2].

The transition from early active to late active represents the point at which patients are detected (for those persons for whom detection does eventually occur) and isolation then occurs from this point forward (i.e. applies during the late disease phase only, see Section 2). This transition point is also intended to represent the point of admission to hospital or transition from hospital ward to intensive care for patients for whom this occurs (see Section 1.4).

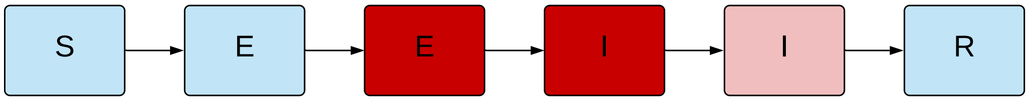

**Figure 1 – Unstratified compartmental model structure.** S = susceptible, E = exposed, I = active disease, R = recovered/removed. Depth of pink/red shading indicates the infectiousness of the compartment.

##### *1.3 Age stratification*

All compartments of this base compartmental structure were stratified by age into five-year bands from 0-4 years of age through to 70-74 years of age, with the final age group being those aged 75 years and older. Heterogeneous baseline contact patterns by age were incorporated using age-specific contact rates estimated by Prem et al. 2017 [3], who combined survey response data with information on national demographic characteristics to produce age-structured mixing matrices with these age groupings. These are then modified by non-pharmaceutical interventions as described in Section 3. Our modelled age groups were chosen to match these mixing matrices. The automatic demographic features of AuTuMN that can be used to simulate births, ageing and deaths were not implemented, because the current questions at hand pertain to the short- to medium-term and the immediate implementation of control strategies, for which population demographics are less relevant.

##### 1.4 Clinical stratification

The age-stratified late exposed/incubation and both the early and late active disease compartments were further stratified into five categories: *1)* asymptomatic, *2)* symptomatic ambulatory, never detected, *3)* symptomatic ambulatory, ever detected, *4)* ever hospitalised, never critical and *5)* ever critically unwell (Figure 2). The proportion of new infectious persons entering stratum 1 (asymptomatic) is age-dependent (as described in Table 4). The proportion of symptomatic patients (strata 2 to 5) ever detected (strata 3 to 5) is set through a parameter that represents the time-varying proportion of all symptomatic patients who are ever detected (the case detection rate, see Section 2). Of those ever symptomatic (strata 2 to 5), a constant but age-specific proportion is considered to be hospitalised (entering strata 4 or 5). Of those hospitalised (entering strata 4 or 5), a fixed proportion was considered to be critically unwell (entering stratum 5, Figure 3).

##### 1.5 Hospitalisation

For COVID-19 patients who are admitted to hospital, the sojourn time in the early and late active compartments is modified, superseding the sojourn times the default values for these compartments, as indicated in Table 3. The point of admission to hospital is considered to be the transition from early to late active disease, such that the sojourn time in the late disease represents the period of time admitted to hospital. For patients admitted to ICU, admission to ICU occurs at this same transition point. For this group, the period of time hospitalised prior to ICU admission is estimated as a proportion of the early active period, such that the early active period represents both the period ambulatory in the community and the period in hospital prior to ICU admission.

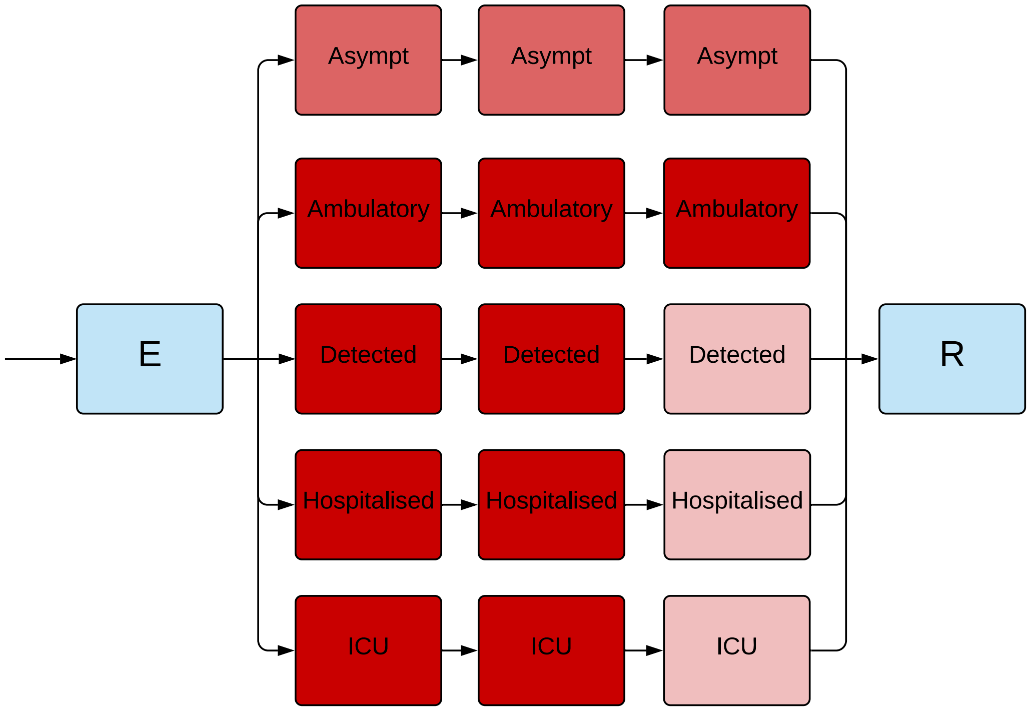

**Figure 2 – Illustration of the implementation of the clinical stratification.** Depth of pink/red shading indicates the infectiousness of the compartment. Typical parameter values presented, although the infectiousness of asymptomatic persons is varied in calibration.

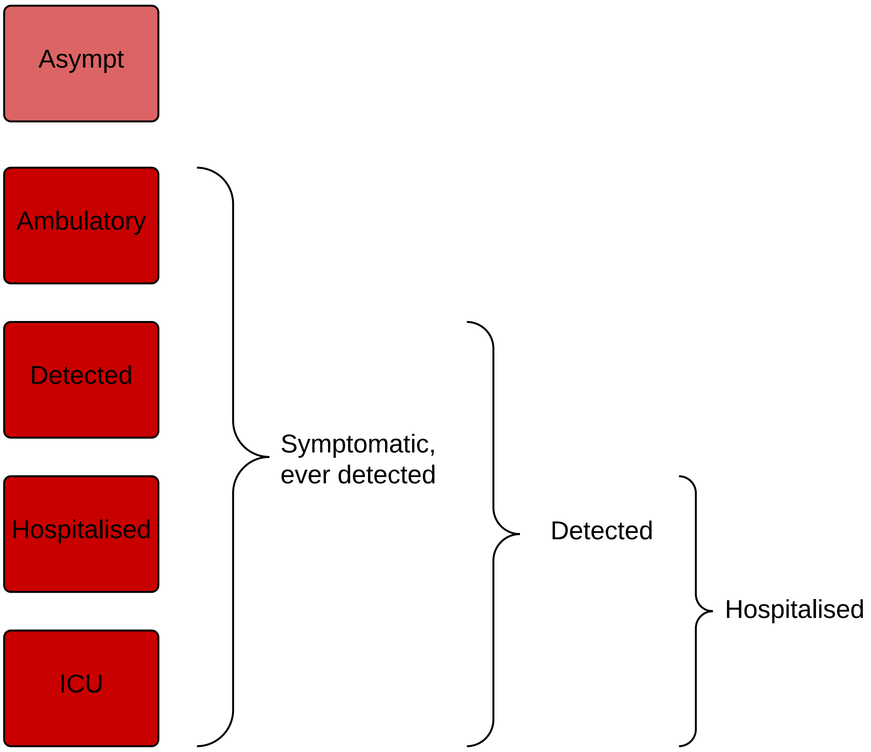

**Figure 3** – Illustration of the rationale for the clinical stratification.

##### 1.6 Infectiousness

Asymptomatic persons are assumed to be less infectious per unit time active than symptomatic persons not undergoing case isolation (typically by around 50%, although this is varied in calibration/uncertainty analysis). Infectiousness is also decreased for persons who have been detected to reflect case isolation, and for those admitted to hospital or ICU to reflect infection control procedures (by 80% for both groups). Presymptomatic individuals are presumed to have equivalent infectiousness to those with early active COVID-19.

##### 1.7 Application of COVID-19-related death

Age-specific infection fatality rates (IFRs) were applied and distributed across strata 4 and 5, with no deaths typically applied to the first three strata. A ceiling of 50% is set on the proportion of those admitted to ICU (entering stratum 5) who die. If the infection fatality rate is greater than this ceiling, the proportion of critically unwell persons dying was set to 50%, with the remainder of the infection fatality rate then applied to the hospitalised proportion. Otherwise, if the infection fatality rate is less than half of the absolute proportion of persons critically unwell, the infection fatality rate is applied entirely through stratum 5 (such that the proportion of critically unwell persons dying in that age group becomes $<$50% and the proportion of stratum 4 dying is set to zero). In the event that the infection fatality rate for an age group is greater than the total proportion hospitalised (which is unusual, but could occur for the oldest age group under certain parameter configurations), the remaining deaths are assigned to the asymptomatic stratum. This approach was chosen because the proportion of persons entering this stratum does not vary over time and the dynamics are therefore identical to assigning the deaths to any of the first three strata, because the sojourn times in the infected compartments are the same for each of these strata. We used the age-specific IFRs previously estimated from age-specific death data from 45 countries and results from national-level seroprevalence surveys [4] as indicated in Table 4. We allowed IFRs to vary around the previously published point estimates in order to incorporate uncertainty and to allow IFRs to vary by country (see Calibration section).

| Clinical stratum | Stratum name | Pre-symptomatic | Early | Late |
|---|---|---|---|---|
| 1 | Asymptomatic | 0.5 | 0.5 | 0.5 |
| 2 | Symptomatic ambulatory never detected | 1 | 1 | 1 |
| 3 | Symptomatic ambulatory ever detected | 1 | 1 | 0.2 |
| 4 | Hospitalised never critical | 1 | 1 | 0.2 |
| 5 | Ever critically unwell | 1 | 1 | 0.2 |

**Table 1 – Illustration of the relative infectiousness of disease compartments by clinical stratification and stage of infection.**

### 2 Case detection rate

#### 2.1 General approach

We calculate a time-varying case detection rate, being the proportion of all symptomatic cases (clinical strata 2 to 5) that are detected (clinical strata 3 to 5). This proportion is informed by the number of tests performed using the following formula:

$$CDR(time) = 1 - e^{-shape \times tests(time)}$$

*time* is the time in days from the 31$^{st}$ December 2019 and *tests(time)* is the number of tests per capita done on that date. To determine the value of the shape parameter, we solve this equation based on the assumption that a certain daily testing rate *tests(time)* is associated with a certain *CDR(time)*. Solving for *shape* yields:

$$shape = \frac{-log(1 - CDR(time))}{tests(time)}$$

That is, if it is assumed that a certain daily per capita testing rate is associated with a certain proportion of symptomatic cases detected, we can determine *shape*. As this relationship is not well understood and unlikely to be consistent across all settings, we vary the *CDR* that is associated with a certain per capita testing rate during uncertainty/calibration. Given that the *CDR* value can be varied widely, the purpose of this is to incorporate changes in the case detection rate that reflect real historical profile in changes in testing capacity over time.

### 3 Implementation of non-pharmaceutical interventions

A major part of the rationale for the development of this model was to capture the past impact of non-pharmaceutical interventions (NPIs) and produce future scenarios projections with the implementation or release of such interventions.

#### 3.1 Isolation and quarantine

For persons who are identified with symptomatic disease and enter clinical stratum 3, self-isolation is assumed to occur and their infectiousness is modified, as described above. The proportion of ambulatory symptomatic persons effectively identified through the public health response by any means is determined by the case detection rate as described above.

#### 3.2 Community quarantine or “lockdown” measures

For all NPIs relating to reduction of human mobility or “lockdown” (i.e. all NPIs other than isolation and quarantine mentioned above), these interventions are implemented through dynamic adjustments to the age-assortative mixing matrix. The baseline mixing matrices of Prem et al. [3] are synthetic and do not represent direct observations or reports from surveys (in the case of the 144 countries to which they were extrapolated from observations in the eight “POLYMOD” countries of Western Europe). Although synthetic, the matrices are contextualised to national demographic information, including country-specific data that include household size, workforce participation and school enrolment. Further, the matrices presented are easily machine-readable and appear to be plausible representations of contact structures within these countries.

The matrices also have the major advantage of allowing for disaggregation of total contact rates by location, i.e. home, work, school and other locations. This disaggregation allows for the simulation of various NPIs in the local context by dynamically varying the contribution of each location to reflect the historical implementation of the interventions.

The corresponding mixing matrix (denoted $C_0$) is presented for each individual model application, using the standard convention that a row represents the average numbers of age-specific contacts per day for a contact recipient of a given age-group. In other words, the element $C_{0i,j}$ is the average number of contacts per day that an individual of age-group $j$ makes with individuals of age-group $i$.

This matrix results from the summation of the four context-specific contact matrices provided by Prem et al.: $C_0 = C_H + C_S + C_W + C_L$, where $C_H$, $C_S$, $C_W$ and $C_L$ are the age-specific contact matrices associated with households, schools, workplaces and other locations, respectively.

In our model, the contributions of the matrices $C_H$, $C_S$, $C_W$ and $C_L$ vary with time such that the input contact matrix can be written:

$$C(t) = h(t)^2 \times C_H + s(t)^2 \times C_S + w(t)^2 \times C_W + l(t)^2 \times C_L$$

The modifying functions are all squared to capture the effect of the mobility changes on both the infector and the infectee in any given interaction that could potentially result in transmission. The modifying functions incorporate functions fitted to Google mobility data, and may additionally incorporate microdistancing effects, depending on the location.

##### 3.3 School closures/re-openings

Reduced attendance at schools is represented through the function $s(t)$, which represents the proportion of all school students currently attending on-site teaching. If schools are fully closed, $s(t) = 0$ and $C_S$ does not contribute to the overall mixing matrix $C(t)$. $s(t)$ is calculated through a series of estimates of the proportion of students attending schools, to which a smoothed step function is fitted. Note that the dramatic changes in this contribution to the mixing matrix with school closures/re-openings is a more dramatic change than is seen with the simulation of policy changes in workplaces and other locations (which are determined by empiric data and so do not vary so abruptly and do not fall to zero).

##### 3.4 Workplace closures

Workplace closures are represented by proportionally reducing the contribution of workplace contacts to the total mixing matrix over time. This is achieved through the scaling function $w(t)$ which modifies the contribution of $C_W$ to the overall mixing matrix $C(t)$. The profile of this function is set by fitting a polynomial spline function to Google mobility data for workplace attendance (Table 2).

##### 3.5 Community-wide movement restriction

Community-wide movement restriction (or "lockdown") measures are represented by proportionally reducing the contribution of the other locations contacts to the total mixing matrix over time. This is achieved through the scaling function $l(t)$ which modifies the contribution of $C_L$ to the overall mixing matrix $C(t)$. The profile of this function is set by fitting a polynomial spline function to an average of Google mobility data for various locations, as indicated in Table 2.

| Prem "location" | Approach | Google mobility types |
|---|---|---|
| School | Policy response | Not applicable |
| Household | Google mobility | Residential |
| Workplace | Google mobility | Workplace |
| Other locations | Google mobility | Unweighted average of:<br>• Retail and recreation<br>• Grocery and pharmacy<br>• Parks<br>• Transit stations |

**Table 2 – Mapping of Google mobility data to contact locations** (as defined by Prem et al.)

##### 3.6 Microdistancing

Interventions other than those that prevent people coming into contact with one another are known to be important to COVID-19 transmission and epidemiology, such as maintaining interpersonal physical distance and the wearing of face coverings. We therefore implemented a "microdistancing" function to represent a reduction in the rate of contact in locations that is not captured through Google mobility data. This microdistancing function reduces the values of all elements of the mixing matrices by a certain proportion. These time-varying functions multiplicatively scale the location-specific contact rate modifiers $h(t)$, $s(t)$, $w(t)$ and $l(t)$.

### 4 Parameters

#### 4.1 Non-age-stratified parameters

| Parameter | Value | Rationale |
|---|---|---|
| Incubation period | Calibration parameter, truncated normal distribution, mean 5.5 days | Estimates of the incubation period have included 5.1 days, 5.2 days and 4.8 days [5] [6] [7] [8]. A systematic review [2] found that data are best fitted by a log-normal distribution (mean 5.8 days, CI 5.0 to 6.7, median 5.1 days). Our systematic review [9] found that estimates of the mean incubation period have varied from 3.6 to 7.4 days. |
| Proportion of incubation period infectious | 50% | Infectiousness is considered to be present throughout a considerable proportion of the incubation period, based on analyses of confirmed source-secondary pairs [10] and early findings that the incubation period was similar to the serial interval [5]. The study of source-secondary pairs was also the primary reference cited by a review of the infectious period that identified studies that quantified the pre-symptomatic period, which concluded that the median pre-symptomatic period could range from less than one to four days [11]. |
| Active period (regardless of detection/isolation, for clinical strata 1 to 3) | Calibration parameter, truncated normal distribution, mean 8 days | This quantity is difficult to estimate, given that identified cases are typically quarantined. Studies in settings of high case ascertainment and an effective public health response have suggested a duration of greater than 5.5 days [8]. PCR positivity, which may continue for up to two to three weeks from the point of symptom onset [10] [11], is difficult to interpret and does not necessarily indicate infectiousness. Consistent with these findings, the duration infectious for asymptomatic persons has been estimated at 6.5 to 9.5 days [11] (although in our model, this would include the pre-symptomatic infectious period). |
| Proportion of infectious period before isolation or hospitalisation can occur | 0.333 | Assumed |

Continuation of Table 3

| **Parameter** | **Value** | **Rationale** |
|---|---|---|
| Disease duration prior to admission for hospitalised patients not critically unwell (i.e. early active sojourn time, stratum 4) | 7.7 days | Mean value from ISARIC cohort, as reported on 4th October 2020 in Table 6 [12], and similar to the expected mean from earlier reports from ISARIC [13]. This cohort represents high-income countries better than low and middle-income countries, with the United Kingdom contributing data on the greatest number of patients, followed by France. Earlier estimates of this quantity from China included 4.4 days [5]. |
| Duration of hospitalisation if not critically unwell (late active sojourn time, stratum 4) | 12.8 days | Mean value from the ISARIC cohort, as reported on 4th October 2020 in Table 6 [12]. |
| ICU duration (late active sojourn time, stratum 5) | 10.5 days | Mean duration of stay in ICU/HDU from ISARIC cohort for patients with complete data, as reported on 10th October 2020 Table 6 [12]. Many other studies reporting on the average duration of ICU stay suffer from right-truncation issues, often estimating 7-10 days length of stay. |
| Duration of time prior to ICU for patients admitted to ICU | 10.5 days | Calculated as the sum of the time from symptom onset to hospital admission (7.7 days above) plus the duration from hospital admission to ICU admission reported by October ISARIC report (2.8 days) [12]. |
| Relative infectiousness of asymptomatic persons (per unit time with active disease) | 0.5 | Assumed |
| Relative infectiousness of persons admitted to hospital or ICU | 0.2 | Assumed |
| Relative infectiousness of identified persons in isolation | 0.2 | Assumed |
| Proportion of hospitalised patients ever admitted to ICU | 0.17 | Assumed |

**Table 3 – Universal (non-age-stratified) model parameters.** Point estimates are used as model parameters except where ranges are indicated in calibration parameter table below in calibration table.

##### 4.2 Age-specific parameters

| Age group (years) | Clinical fraction[a] | Relative susceptibility to infection | Infection fatality rate | Proportion of symptomatic patients hospitalised |
|---|---|---|---|---|
| 0 to 4 | 0.29 | 0.36 | $3 \times 10^{-5}$ | 0.0777 |
| 5 to 9 | 0.29 | 0.36 | $1 \times 10^{-5}$ | 0.0069 |
| 10 to 14 | 0.21 | 0.36 | $1 \times 10^{-5}$ | 0.0034 |
| 15 to 19 | 0.21 | 1 | $3 \times 10^{-5}$ | 0.0051 |
| 20 to 24 | 0.27 | 1 | $6 \times 10^{-5}$ | 0.0068 |
| 25 to 29 | 0.27 | 1 | $1.3 \times 10^{-4}$ | 0.0080 |
| 30 to 34 | 0.33 | 1 | $2.4 \times 10^{-4}$ | 0.0124 |
| 35 to 39 | 0.33 | 1 | $4.0 \times 10^{-4}$ | 0.0129 |
| 40 to 44 | 0.40 | 1 | $7.5 \times 10^{-4}$ | 0.0190 |
| 45 to 49 | 0.40 | 1 | $1.21 \times 10^{-3}$ | 0.0331 |
| 50 to 54 | 0.49 | 1 | $2.07 \times 10^{-3}$ | 0.0383 |
| 55 to 59 | 0.49 | 1 | $3.23 \times 10^{-3}$ | 0.0579 |
| 60 to 64 | 0.63 | 1 | $4.56 \times 10^{-3}$ | 0.0617 |
| 65 to 69 | 0.63 | 1.41 | $1.075 \times 10^{-2}$ | 0.1030 |
| 70 to 74 | 0.69 | 1.41 | $1.674 \times 10^{-2}$ | 1.072 |
| 75 and above | 0.69 | 1.41 | $5.748 \times 10^{-2}$ [b] | 0.0703 |
| Source/ rationale | Model fitting to age-distribution of early cases in China, Italy, Japan, Singapore, South Korea and Canada taken from upper-left panel of Figure 2b of [14]. | Conversion of odds ratios presented in Table S15 of Zhang et al. 2020 to relative risks using data presented in Table S14 of the same study [15].[c] | Estimated from pooled analysis of data from 45 countries from Table S3 of O'Driscoll et al [4]. Values consistent with previous estimates using serosurveys performed in Spain [16]. | Estimates from the Netherlands as the first wave of infections declined from 4th May to 21st July [17]. |

**Table 4 – Age-stratified parameter values.** Age-stratified parameters not varied during calibration, or varied through a common adjuster parameter.
[a] Proportion of incident cases developing symptoms.
[b] Weighted average of IFR estimates for 70 to 79 and 80 and above age groups.
[c] Note the relative magnitude of these values are similar to those estimated by the analysis we use to estimate the age-specific clinical fraction.[14]

### 5 Calculation of outputs

#### 5.1 Incidence

Incidence is calculated as any transitions into the early active compartment (*"I"*).

#### 5.2 Hospital occupancy

This is calculated as the sum of three quantities:

1. All persons in the late active compartment in clinical stratum 4, representing those admitted to hospital but never critically unwell.

2. All persons in the late active compartment in clinical stratum 5, representing those currently admitted to ICU.

3. A proportion of the early active compartment in clinical stratum 5, representing those who will be admitted to ICU at a time in the future. This proportion is calculated as the quotient of *1)* the difference between the pre-ICU period and the pre-hospital period for clinical stratum 4, divided by *2)* the total pre-ICU period. That is, a proportion of the pre-ICU period is considered to represent patients in hospital who have not yet been admitted to ICU.

#### 5.3 ICU occupancy

This is calculated as all persons in the late active compartment in clinical stratum 4.

#### 5.4 Seropositive proportion

This is calculated as the proportion of the population in the recovered (*"R"*) compartment.

#### 5.5 COVID-19-related mortality

This is calculated as all transitions representing death, exiting the model. This is implemented as depletion of the late active compartment.

##### 5.6 Notifications

Local case notifications are calculated as transitions from the early to the late active compartment for clinical strata 3 to 5. Imported case notifications are calculated as the product of the time-variant importation rate (which has been inflated for incomplete case detection) and the case detection rate.

#### 6 Calibration

##### 6.1 General approach

The model was calibrated using an adaptive Metropolis (AM) algorithm. In particular, we used the algorithm based on adaptive Gaussian proposal functions proposed by Haario *et al.* to sample parameters from their posterior distributions [18]. We ran seven independent AM chains and combined the samples of the seven chains to project epidemic trajectories over time. The definitions of the prior distributions and the likelihood are detailed as follows.

##### 6.2 Likelihood function

Likelihood functions are derived from comparing model outputs to target data at each time point nominated for calibration. The dispersion parameters of these distribution are included as calibration parameters and varied during the calibration approach to improve calibration efficiency.

##### 6.3 Variation of infection fatality rate and symptomatic proportions

Whether age-specific infection fatality rates (IFRs) are significantly different in low- and middle-income settings from those in high-income settings remains highly uncertain. For this application to the Philippines, we adjust the IFRs described above according to a factor that modifies the age-specific IFR for each age group relative to the baseline values described in the previous section, allowing them to vary from those reported up to two-fold those values. The age-specific IFRs used in the model are obtained from:

$$IRF_i^* = \frac{IFR_i \times \omega}{IFR_i(\omega - 1) + 1},$$

where $IRF_i^*$ is the modelled IFR for age group $i$, $IFR_i$ is the point estimate reported by O'Driscoll *et al.* for the IFR of age group $i$ [4], and $\omega$ is the uncertainty adjuster varied during model calibration.

Similarly, we incorporated uncertainty around the age-specific proportions of symptomatic individuals by applying an uncertainty adjuster:

$$s_i^* = \frac{s_i \times \gamma}{s_i(\gamma - 1) + 1},$$

where $s_i^*$ is the modelled symptomatic proportion for age group $i$, $s_i$ is the point estimate reported by Davies *et al.* for the IFR of age group $i$ [14], and $\gamma$ is the associated uncertainty adjuster varied during model calibration.

##### 6.4 Calibration parameters

| **Parameter name** | **Distribution type** | **Distribution parameters** |
|---|---|---|
| Incubation period | Truncated normal | Mean 5.5 days, standard deviation 0.97 days, truncation <1 day |
| Infectious period (for clinical strata 1 to 3) | Truncated normal | Mean 6.5 days, standard deviation 0.77 days, truncation <4 days |
| Risk of infection per contact | Uniform | 0.03 to 0.05 |
| Infection fatality rate adjuster ($\omega$) | Uniform | Range 1.8 to 2.28 |

Continuation of Table 5

| Parameter name | Distribution type | Distribution parameters |
|---|---|---|
| Proportion of symptomatic cases that would be detected with a daily per capita testing rate of one test per ten thousand population | Uniform | Range 0.02 to 0.15 |
| Infectious seed | Uniform | Range 10 to 100 |
| Maximal effect of Minimum Health Standards | Uniform | 0.1 to 0.6 |
| Adjuster applied to age-specific proportion of infections leading to symptoms ("Clinical fraction") | Truncated normal | Mean 1, standard deviation 0.2, truncation $<$0.5 |

**Table 5 – Epidemiological calibration parameters.**

##### 6.5 Calibration targets

We calibrated each of the Philippines models to the daily notification rate (with seven-day moving average smoothing) observed nationally or for the sub-region simulated over time.

For ICU occupancy, we only considered the most recent estimate of ICU occupancy and did not calibrate to multiple time points for this indicator. Similarly for cumulative infection-related deaths, we only calibrated to the most recent data time point available.

tabularx

### 7 Ordinary differential equations

For the clearest description of the model, we refer the reader to our code repository, because our object-oriented approach to software development is intended to be highly transparent and readable. For those who prefer dynamical systems such as this presented in the form of ordinary differential equations, we present the following.

$$\frac{dS_a}{dt} = -\lambda_a(t) \times \sigma_a \times S_a$$
$$\frac{dE_a}{dt} = \lambda_a(t) \times \sigma_a \times S_a - \alpha E_a$$
$$\frac{dP_{a,c}}{dt} = p_{a,c}(t) \times \alpha E_a - \nu P_{a,c}$$
$$\frac{dI_{a,c}}{dt} = \nu P_{a,c} - \gamma_c I_{a,c}$$
$$\frac{dL_{a,c}}{dt} = \gamma_c I_{a,c} - \delta_{a,c} L_{a,c} - \mu_{a,c} L_{a,c}$$
$$\frac{dR_a}{dt} = \sum_c \delta_{a,c} L_{a,c}$$

where

$$\lambda_a = \beta \left[ \sum_{j,c} \frac{\varepsilon \times P_j}{N_j} \times C_{a,j}(t) + \sum_{j,c} \frac{I_{j,c} \times \iota_c + L_{j,c} \times \kappa_c}{N_j} \times C_{a,j}(t) \right]$$
$$\sum_c p_{a,c}(t) = 1, \forall t \in \mathbb{R}$$
$$\mathbf{C}_0 = \mathbf{C}_H + \mathbf{C}_S + \mathbf{C}_W + \mathbf{C}_L$$
$$\mathbf{C}(t) = h(t) \times \mathbf{C}_H + s(t) \times \mathbf{C}_S + w(t) \times \mathbf{C}_W + l(t) \times \mathbf{C}_L$$
$$l(t) = \frac{re(t) + gr(t) + pa(t) + tr(t)}{4}$$

| **Symbol** | **Explanation** |
|---|---|
| $S$ | Persons susceptible to infection |
| $E$ | Persons in the non-infectious incubation period |
| $P$ | Persons in the incubation period |
| $I$ | Persons in the early active disease period, before isolation or hospitalisation may occur |
| $L$ | Persons in the late active disease period, after isolation or hospitalisation may have occurred |
| $R$ | Persons in the recovered period, from which re-infection cannot occur |

| **Symbol** | **Explanation** |
|---|---|
| $t$ | Time |
| $a$ | Compartment of age group a |
| $c$ | Compartment of clinical stratification c |
| $\sigma$ | Relative susceptibility to infection |
| $\alpha$ | Rate of progression from non-infectious to infectious incubation period |
| $\nu$ | Rate of progression from infectious incubation to early active disease |
| $\gamma$ | Rate of progression from early active disease to late active disease |
| $\mu$ | Rate of disease-related death |
| $\varepsilon$ | Relative infectiousness of pre-symptomatic compartment |
| $\iota$ | Clinical stratification infectiousness vector for early active compartment |
| $\kappa$ | Clinical stratification infectiousness vector for late active compartments |
| $\beta$ | Probability of infection per contact between an infectious and susceptible individual |
| $j$ | Infectious populations |
| $p$ | Proportion progressing to each clinical stratification |

| **Symbol** | **Explanation** |
|---|---|
| **C** | Mixing matrix |
| **H** | Household contribution to mixing matrix |
| **W** | Workplace contribution to mixing matrix |
| **O** | Other locations contribution to mixing matrix |
| **S** | Schools contribution to mixing matrix |
| $l$ | Other locations macrodistancing function of time |
| $w$ | Function fit to Google mobility data for workplaces |
| $s$ | Function fit to Google mobility data for schools |
| $re$ | Function fit to Google mobility data for retail and recreation |
| $gr$ | Function fit to Google mobility data for grocery and pharmacy |
| $pa$ | Function fit to Google mobility data for parks |
| $tr$ | Function fit to Google mobility data for transit stations |

### 8 Supplemental figures and tables to main text

#### 8.1 Supplemental Tables

**Table 6 – Laboratory testing facilities by region.**

| Facility Name | Region |
|---|---|
| Batangas Medical Center GeneXpert Laboratory | Calabarzon |
| Daniel O. Mercado Medical Center | Calabarzon |
| De La Salle Medical and Health Sciences Institute | Calabarzon |
| Greencity Medical Center | Calabarzon |
| Lucena United Doctors Hospital and Medical Center | Calabarzon |
| Mary Mediatrix Medical Center | Calabarzon |
| Ospital ng Imus | Calabarzon |
| Qualimed Hospital Sta. Rosa | Calabarzon |
| San Pablo College Medical Center | Calabarzon |
| San Pablo District Hospital | Calabarzon |
| UPLB Covid-19 Molecular Laboratory | Calabarzon |
| Allegiant Regional Care Hospital | Central Visayas |
| Cebu Doctors University Hospital Inc | Central Visayas |
| CebuTBReferenceLaboratory-MolecularFacilityforCOVID-19Testing | Central Visayas |
| Chong Hua Hospital | Central Visayas |
| Governor Celestino Gallares Memorial Medical Center | Central Visayas |

| Facility Name | Region |
|---|---|
| Prime Care Alpha Covid-19 Testing Laboratory | Central Visayas |
| University of Cebu Medical Center | Central Visayas |
| Vicente Sotto Memorial Medical Center (VSMMC) | Central Visayas |
| Amang Rodriguez Memorial Center GeneXpert Laboratory | National Capital Region |
| Asian Hospital and Medical Center | National Capital Region |
| Chinese General Hospital | National Capital Region |
| De Los Santos Medical Center | National Capital Region |
| Dr. Jose N. Rodriguez Memorial Hospital and Sanitarium (TALA) GeneXpert Laboratory | National Capital Region |
| Dr. Jose N. Rodriguez Memorial Hospital and Sanitarium (TALA) RT PCR | National Capital Region |
| Fe del Mundo Medical center | National Capital Region |
| Hi-Precision Diagnostics (QC) | National Capital Region |
| Lung Center of the Philippines (LCP) | National Capital Region |
| Lung Center of the Philippines GeneXpert Laboratory | National Capital Region |
| Makati Medical Center (MMC) | National Capital Region |
| Marikina Molecular Diagnostics Laboratory (MMDL) | National Capital Region |
| National Kidney and Transplant Institute | National Capital Region |
| National Kidney and Transplant Institute GeneXpert Laboratory | National Capital Region |
| Philippine Children's Medical Center | National Capital Region |

| Facility Name | Region |
|---|---|
| Philippine Heart Center GeneXpert Laboratory | National Capital Region |
| SafeguardDNADiagnosticsInc | National Capital Region |
| San Miguel Foundation Testing Laboratory | National Capital Region |
| Singapore Diagnostics | National Capital Region |
| St. Luke's Medical Center - BGC (HB) GeneXpert Laboratory | National Capital Region |
| St. Luke's Medical Center - BGC (SLMC-BGC) | National Capital Region |
| St. Luke's Medical Center - Quezon City (SLMC-QC) | National Capital Region |
| Sta. Ana Hospital - Closed System Molecular Laboratory (GeneXpert) | National Capital Region |
| The Medical City (TMC) | National Capital Region |
| Tondo Medical Center GeneXpert Laboratory | National Capital Region |
| Tropical Disease Foundation | National Capital Region |
| University of Perpetual Help DALTA Medical Center Inc | National Capital Region |
| UP-PGH Molecular Laboratory | National Capital Region |
| UP National Institutes of Health (UP-NIH) | National Capital Region |
| UP Philippine Genome Center | National Capital Region |
| Veteran Memorial Medical Center | National Capital Region |
| Victoriano Luna - AFRIMS | National Capital Region |

#### 8.2 Supplemental Figures

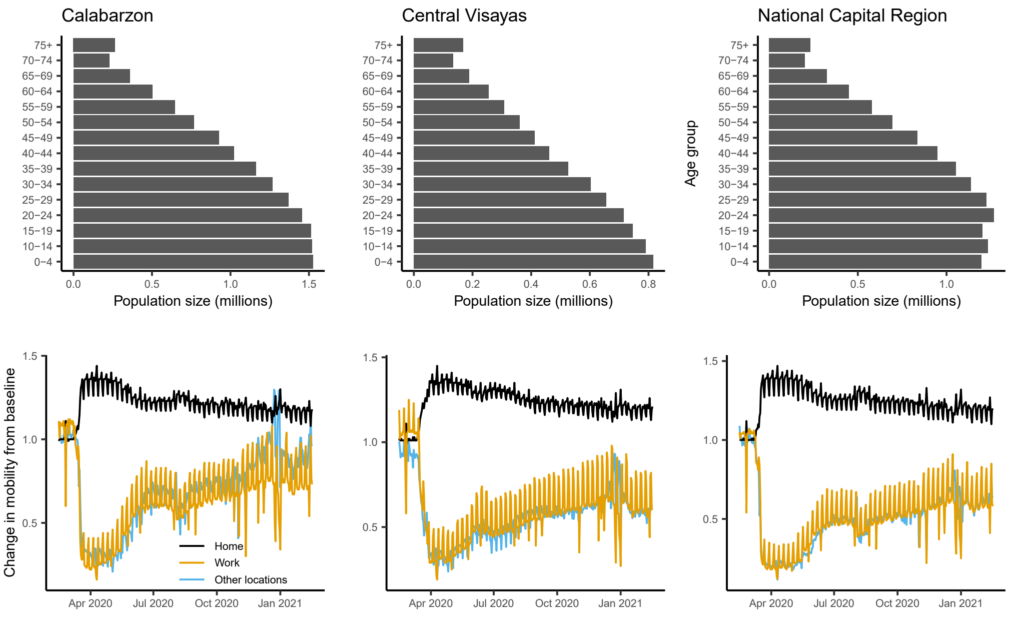

**Figure 4 – Population size and mobility included in the age-structured COVID-19 for three regions of the Philippines. Starting population age distribution (top row) and community quarantine driven mobility adjustments applied to the mixing matrices (bottom row) for Calabarzon (left), Central Visayas (middle), and the National Capital Region (right). Other locations in the mobility plots include retail and recreation, supermarket and pharmacy, parks, and public transport.**

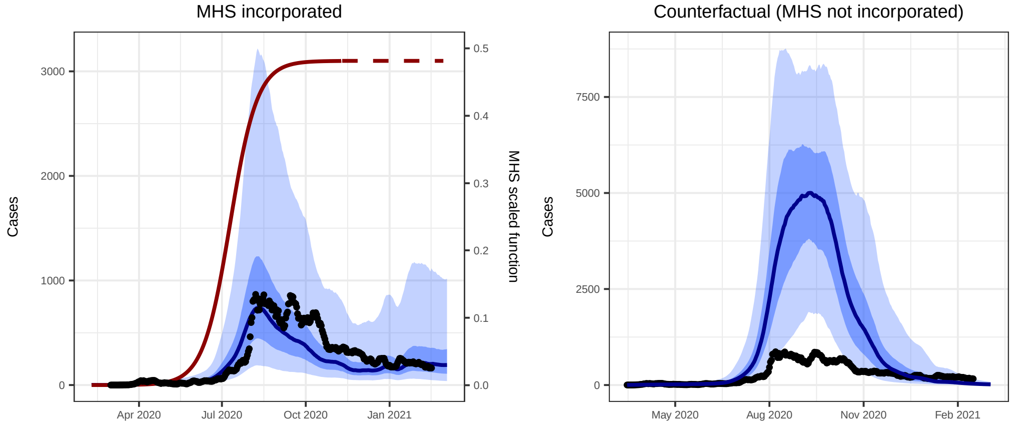

**Figure 5 – Comparison of models that included and did not include Minimum Health Standards (MHS) with daily confirmed cases for Calabarzon. We calibrated the Calabarzon model to daily confirmed cases (black dots; same in both plots with different y-axes), which included MHS (left) and ran a counterfactual scenario that did not include MHS (right). Each plot shows the median modeled detected cases (blue line) with shaded areas representing the 25th to 75th centile (dark blue) and 2·5th to 97·5th centile (light blue) of estimated detected cases. The red curve represents the effect of MHS (i.e., reduced transmission risk per contact) in the model through time. The MHS effect value is squared in the model to account for the reduction in the probability of an infected person passing on the infection and the probability of a contact being infected, prior to adjustment of each cell of the mixing matrix.**

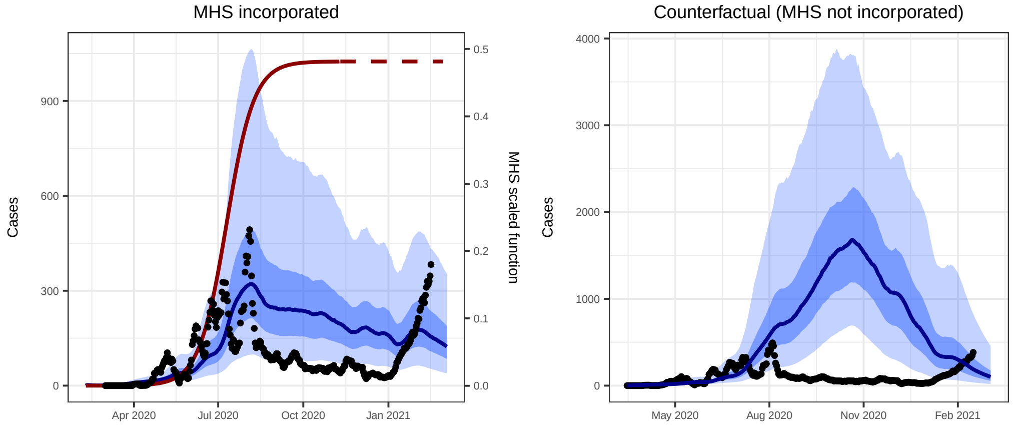

**Figure 6 – Comparison of models that included and did not include Minimum Health Standards (MHS) with daily confirmed cases for Central Visayas. We calibrated the Central Visayas model to daily confirmed cases (black dots; same in both plots with different y-axes), which included MHS (left) and ran a counterfactual scenario that did not include MHS (right). Each plot shows the median modeled detected cases (blue line) with shaded areas representing the 25th to 75th centile (dark blue) and 2·5th to 97·5th centile (light blue) of estimated detected cases. The red curve represents the effect of MHS (i.e., reduced transmission risk per contact) in the model through time. The MHS effect value is squared in the model to account for the reduction in the probability of an infected person passing on the infection and the probability of a contact being infected, prior to adjustment of each cell of the mixing matrix. The recent uptick in cases could be due to a new SARS-CoV-2 Variant of Concern.**

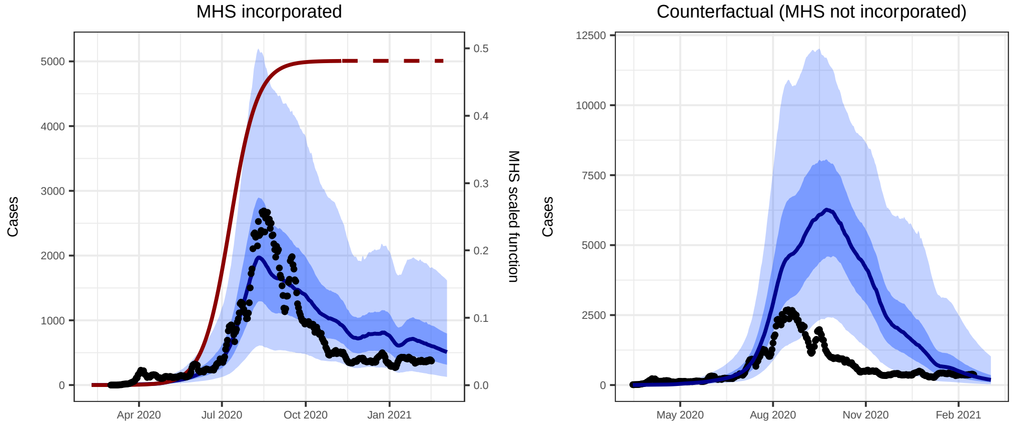

**Figure 7 – Comparison of models that included and did not include Minimum Health Standards (MHS) with daily confirmed cases for the National Capital Region. We calibrated the National Capital Region model to daily confirmed cases (black dots; same in both plots with different y-axes), which included MHS (left) and ran a counterfactual scenario that did not include MHS (right). Each plot shows the median modeled detected cases (blue line) with shaded areas representing the 25th to 75th centile (dark blue) and 2·5th to 97·5th centile (light blue) of estimated detected cases. The red curve represents the effect of MHS (i.e., reduced transmission risk per contact) in the model through time. The MHS effect value is squared in the model to account for the reduction in the probability of an infected person passing on the infection and the probability of a contact being infected, prior to adjustment of each cell of the mixing matrix.**

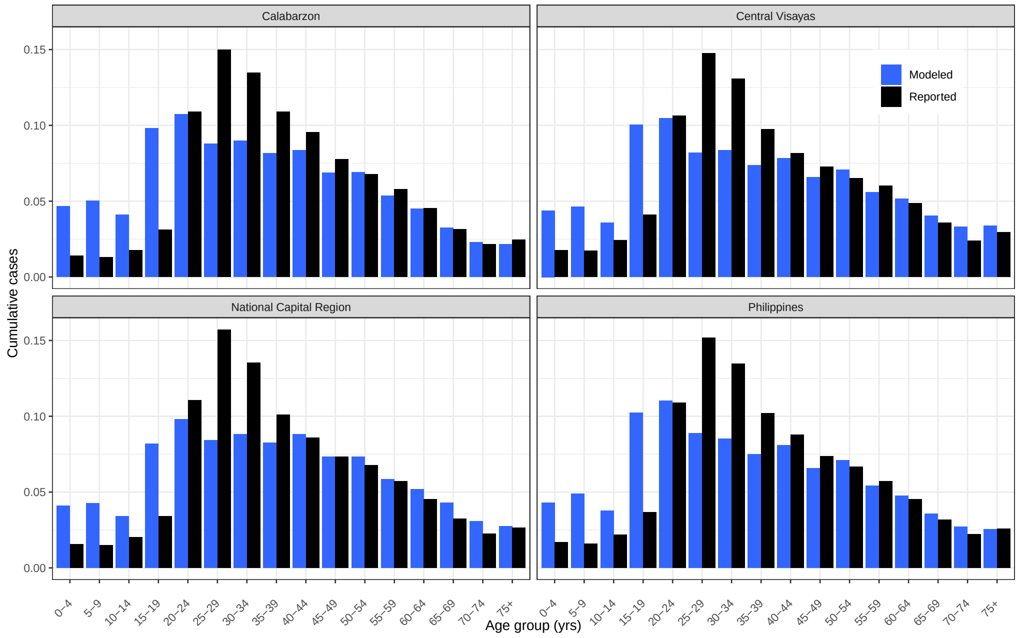

**Figure 8 – Proportional modeled (blue bars) versus reported (black bars) cumulative cases by age group. The model over predicted cumulative cases for young age groups and under predicted cumulative cases for middle aged groups, with good fit for age groups over 40 years. There were 2,174 reported cases excluded from these plots because there was age recorded.**

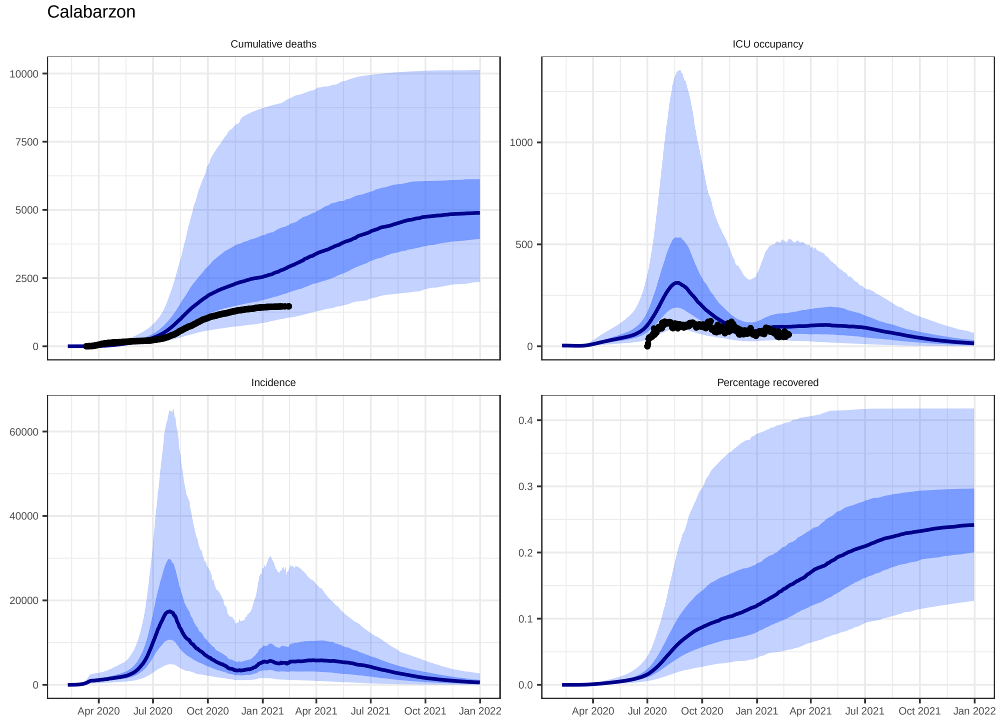

**Figure 9 – Model estimated epidemic indices from the calibrated Calabarzon model. Modeled median cumulative deaths, ICU occupancy, incidence, and percentage of the population recovered from COVID-19 (blue line) with shaded areas for 25th to 75th centile (dark blue) and 2·5th to 97·5th centile (light blue) and overlaid with reported cumulative deaths and ICU occupancy (black dots).**

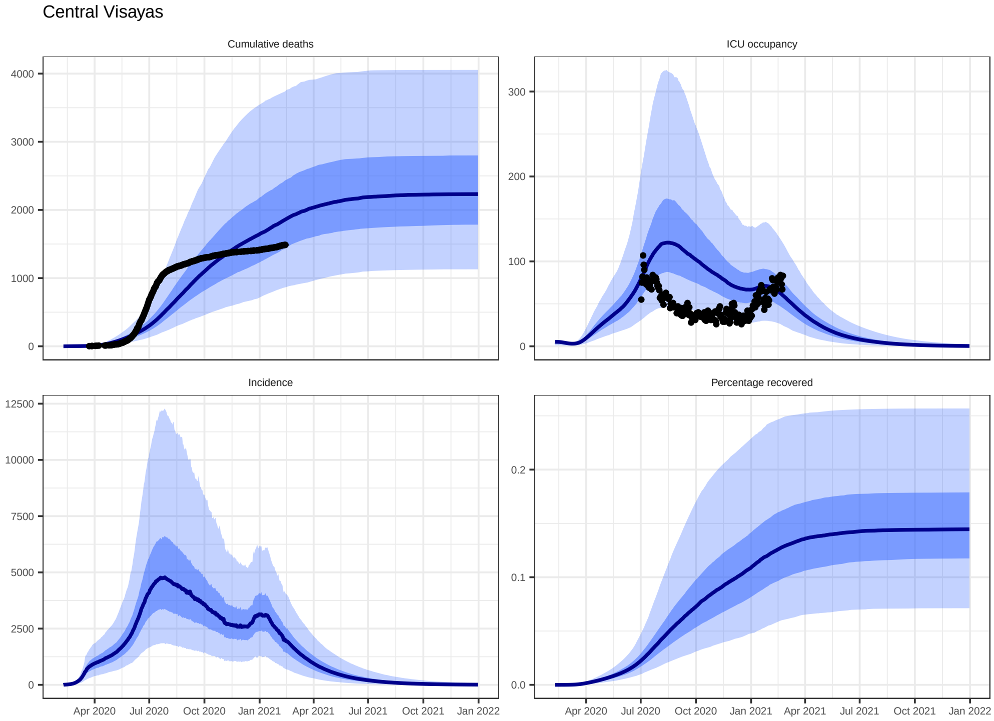

**Figure 10 – Model estimated epidemic indices from the calibrated Central Visayas model. Modeled median cumulative deaths, ICU occupancy, incidence, and percentage of the population recovered from COVID-19 (blue line) with shaded areas for 25th to 75th centile (dark blue) and 2·5th to 97·5th centile (light blue) and overlaid with reported cumulative deaths and ICU occupancy (black dots).**

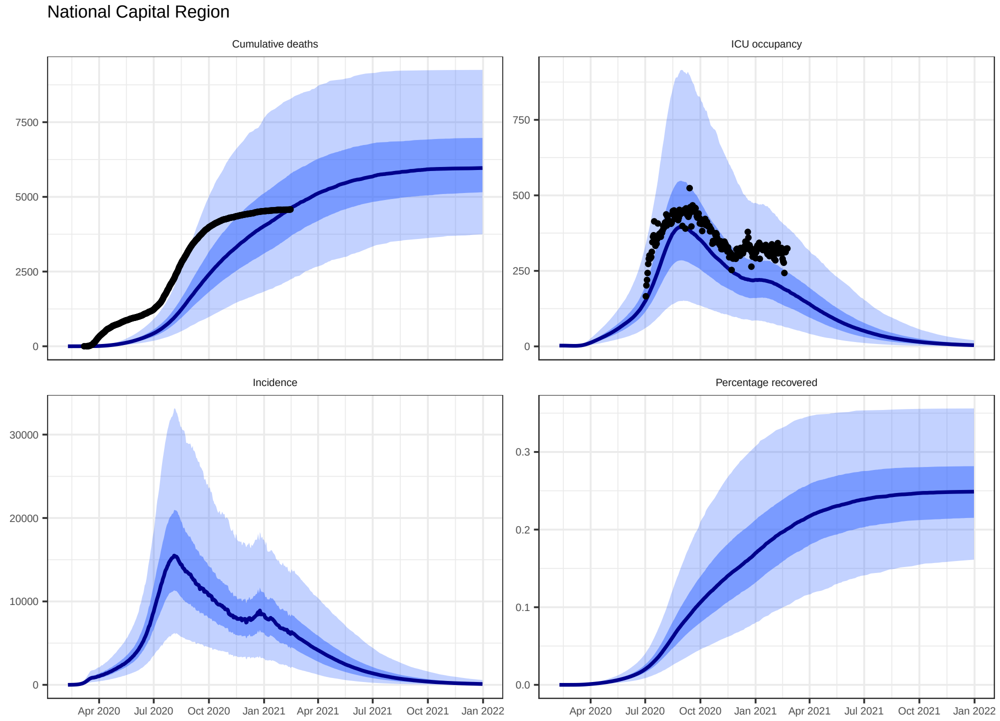

**Figure 11 – Model estimated epidemic indices from the calibrated National Capital Region model. Modeled median cumulative deaths, ICU occupancy, incidence, and percentage of the population recovered from COVID-19 (blue line) with shaded areas for 25th to 75th centile (dark blue) and 2·5th to 97·5th centile (light blue) and overlaid with reported cumulative deaths and ICU occupancy (black dots).**

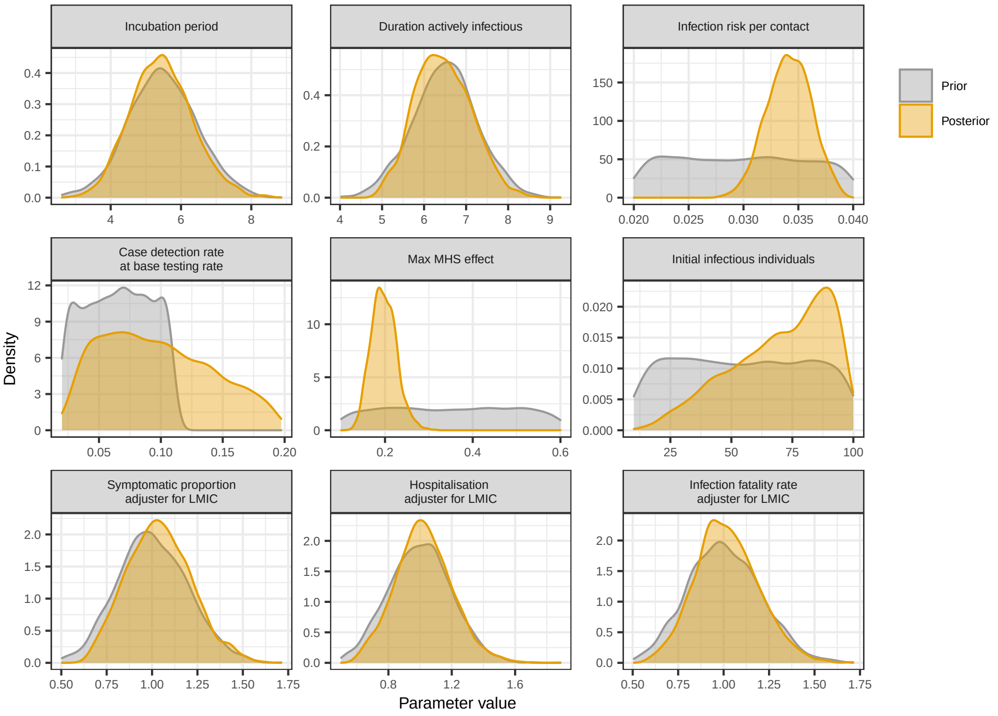

**Figure 12 – Histograms of prior and posterior epidemiological parameter distributions for the Philippines national model. MHS refers to Minimum Health Standards. All parameters with the term "adjuster" allow for modification to the best estimates from the literature (i.e., the priors). The parameter value of the posterior provides the odds ratio used in the model to adjust the odds of an event. For example, "infection fatality rate" was adjusted downwards in the majority of the posterior probability distribution, but adjusted upwards as much as 1·5-1·75 fold in a small minority.**

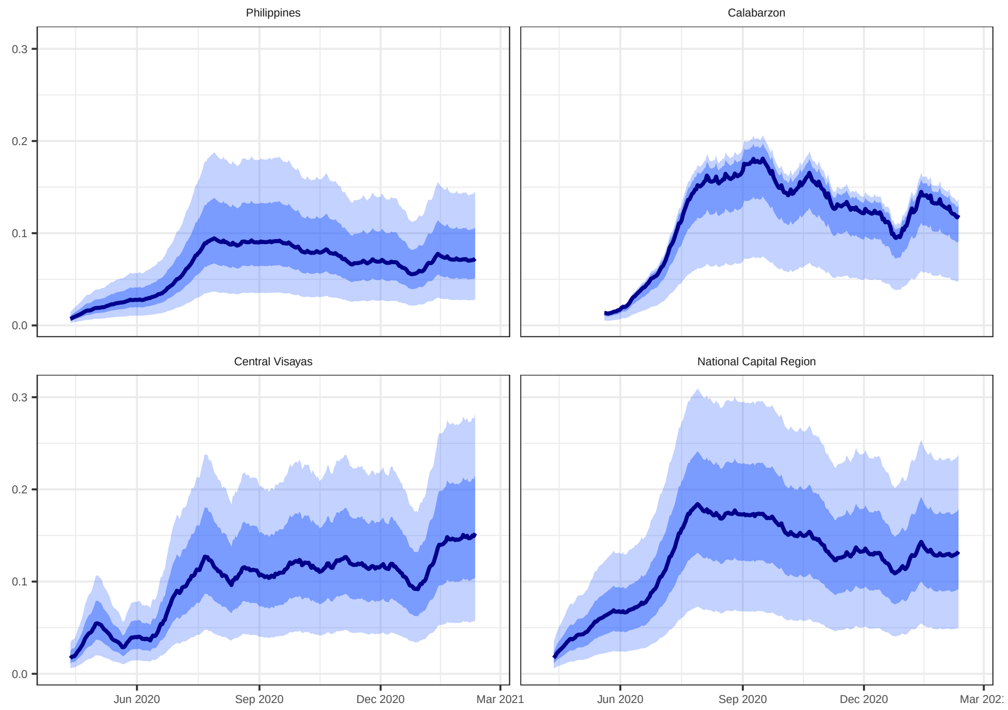

**Figure 13 – Model estimated case detection rates varied across regions and through time, peaking at ¡50%. We derived values for the symptomatic cases detected through time from the daily number of tests performed, the population size, and the calibrated parameter representing the value of the case detection rate given a testing rate of one test per 10,000 persons per day. We provide a list of laboratory facilities that conducted tests in the three regional models in Table S1.**

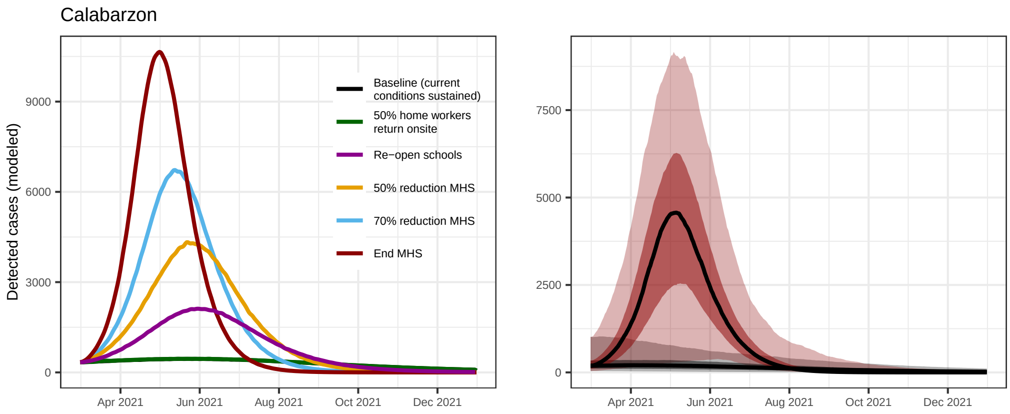

**Figure 14 – Epidemic scenario projections for Calabarzon. Median estimates of daily confirmed cases expected under different policy changes (left). Median estimates of daily confirmed cases (lines) with 25th to 75th centiles (dark shading) and 2·5th to 97·5th centiles (light shading) for the baseline scenario (where current conditions are carried forward) and for the scenario where MHS policy ends. Note the different y-axes on each plot.**

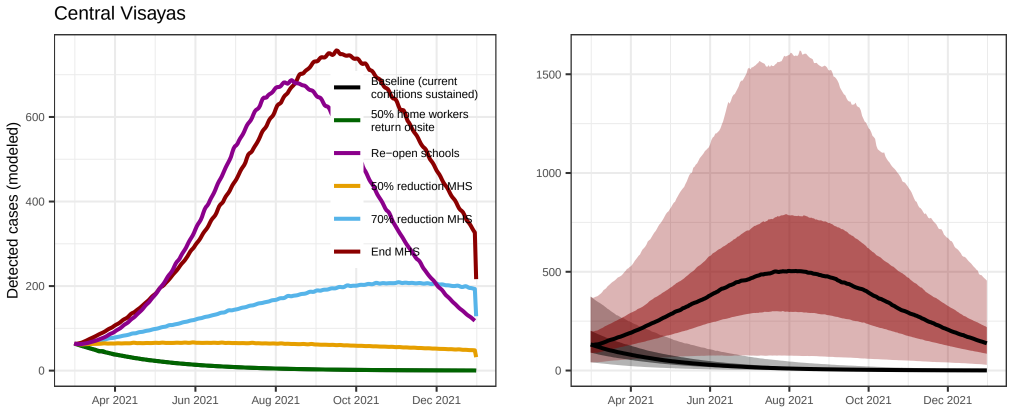

**Figure 15 – Epidemic scenario projections for Central Visayas. Median estimates of daily confirmed cases expected under different policy changes (left). Median estimates of daily confirmed cases (lines) with 25th to 75th centiles (dark shading) and 2·5th to 97·5th centiles (light shading) for the baseline scenario (where current conditions are carried forward) and for the scenario where MHS policy ends. Note the different y-axes on each plot.**

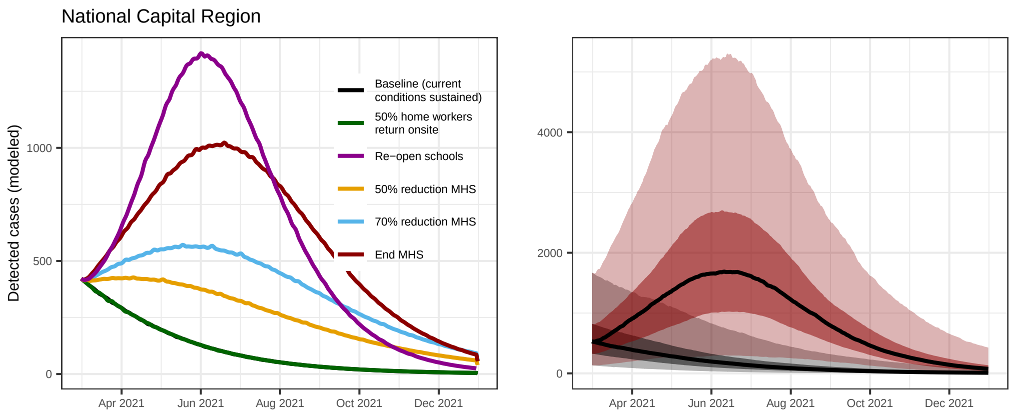

**Figure 16 – Epidemic scenario projections for the National Capital Region. Median estimates of daily confirmed cases expected under different policy changes (left). Median estimates of daily confirmed cases (lines) with 25th to 75th centiles (dark shading) and 2·5th to 97·5th centiles (light shading) for the baseline scenario (where current conditions are carried forward) and for the scenario where MHS policy ends. Note the different y-axes on each plot.**

#### References

[1] J. M. Trauer et al. Modular programming for tuberculosis control, the "AuTuMN" platform. *BMC Infectious Diseases*, 17(1):546, dec 2017.

[2] C. G. McAloon et al. The incubation period of COVID-19: A rapid systematic review and meta-analysis of observational research. Technical Report 8, aug 2020.

[3] K. Prem et al. Projecting social contact matrices in 152 countries using contact surveys and demographic data. *PLoS Computational Biology*, 13(9):e1005697, sep 2017.

[4] M. O'Driscoll et al. Age-specific mortality and immunity patterns of SARS-CoV-2. *Nature*, nov 2020.

[5] J. Zhang et al. Evolving epidemiology and transmission dynamics of coronavirus disease 2019 outside Hubei province, China: a descriptive and modelling study. *The Lancet Infectious Diseases*, 20(7), 2020.

[6] S. A. Lauer et al. The Incubation Period of Coronavirus Disease 2019 (COVID-19) From Publicly Reported Confirmed Cases: Estimation and Application. *Annals of Internal Medicine*, 172(9):577–582, may 2020.

[7] Q. Li et al. Early transmission dynamics in Wuhan, China, of novel coronavirus-infected pneumonia, mar 2020.

[8] Q. Bi et al. Epidemiology and Transmission of COVID-19 in Shenzhen China: Analysis of 391 cases and 1,286 of their close contacts. *medRxiv*, pp. 2020.03.03.20028423, mar 2020.

[9] M. T. Meehan et al. Modelling insights into the COVID-19 pandemic, jun 2020.

[10] X. He et al. Temporal dynamics in viral shedding and transmissibility of COVID-19. *Nature Medicine*, pp. 2020.03.15.20036707, mar 2020.

[11] A. W. Byrne et al. Inferred duration of infectious period of SARS-CoV-2: rapid scoping review and analysis of available evidence for asymptomatic and symptomatic COVID-19 cases. *BMJ open*, 10(8):e039856, aug 2020.

[12] M. Pritchard et al. ISARIC Clinical Data Report 4 October 2020. *medRxiv*, pp. 2020.07.17.20155218, jan 2020.

[13] ISARIC. ISARIC (International Severe Acute Respiratory and Emerging Infections Consortium) COVID-19 Report: 08 June 2020. Technical report, 2020.

[14] N. G. Davies et al. Age-dependent effects in the transmission and control of COVID-19 epidemics. *Nature Medicine*, pp. 2020.03.24.20043018, may 2020.

[15] J. Zhang et al. Changes in contact patterns shape the dynamics of the COVID-19 outbreak in China. *Science*, 368(6498):1481–1486, jun 2020.

[16] M. Pollán et al. Prevalence of SARS-CoV-2 in Spain (ENE-COVID): a nationwide, population-based seroepidemiological study. *Lancet (London, England)*, 0(0), jul 2020.

[17] Epidemiologische situatie COVID-19 in Nederland. Technical report.

[18] H. Haario et al. An adaptive Metropolis algorithm. *Bernoulli*, 2001.
